# Variantscape: Retrospective Study of Large Language Model-Based Extraction of Variant-Treatment-Cancer Associations from Published Oncology Abstracts

**DOI:** 10.64898/2026.08.02.26359492

**Authors:** Marie Wosny, Andrei Stefan Blindu, Maximilian Boesch, Tobias Peres, Thibault Niederhauser, Martin Früh, Christian Rothermundt, Janna Hastings

## Abstract

**Objectives:** Precision oncology relies on accurate interpretation of tumour-detected gene variants, to guide personalised treatment decisions. However, interpretation of variants in context requires extensive information that is often buried within unstructured biomedical literature and obscured by inconsistent nomenclature, making manual retrieval labour-intensive and prone to omissions. In this study, our aim was to evaluate the potential of large language models to extract variant-treatment-cancer associations at scale from published abstracts.

**Design:** We developed *Variantscape*, a large-scale, automated pipeline and open-access web tool. It integrates traditional natural language processing methods with state-of-the-art large language models to extract, standardise, and analyse co-associations between genetic variants, cancer types, and therapeutic interventions from published biomedical abstracts.

**Results:** From over 3.7 million abstracts published between January 2014 and October 2026, 886,195 articles containing gene names were eligible for extraction. Among these, 37,895 (4.3%) mentioned a variant, cancer type, and therapeutic agent together, covering 11,985 unique variants across 197 cancer types and 547 therapeutic agents. LLM verification against the abstract text confirmed at least one association in 35,224 papers, giving a literature-derived graph of 10,571 nodes and 44,644 verified edges. Canonical alterations in well-studied cancers (e.g., *BRAF* V600E in melanoma) were strongly linked to established treatments, while several rare variants also emerged with high-confidence literature support.

**Conclusions:** By applying large language models to biomedical literature, *Variantscape* enables scalable, context-aware extraction of trilateral variant-treatment-cancer relationships. This approach supports early evidence synthesis/hypothesis generation, highlights underrecognized or rare associations for MTB preparation, and offers a practical resource for accelerating discovery and supporting precision oncology research and translation.

**Funding:** This study was funded by the School of Medicine at the University of St. Gallen in Switzerland under grant number 2300380.

**Article Summary:** *Strengths and limitations of this study:*

● *Variantscape* combines traditional natural language processing with large language models to enable scalable, automated extraction of variant-cancer-treatment relationships from over 3.7 million biomedical abstracts, substantially reducing the manual search burden inherent to existing molecular tumour board workflows.
● The pipeline applies large language model-based inference to standardise inconsistent genetic nomenclature across the literature, improving the comparability and reliability of extracted associations that would otherwise be obscured by terminological variation.
● *Variantscape* is designed as a continuously updatable, open-access resource, meaning it can incorporate newly published evidence without requiring repeated manual intervention.
● The pipeline relies solely on abstract-level text rather than full-text articles, which may limit the depth and granularity of extracted associations, as key methodological details, variant context, and nuanced clinical findings are frequently reported only within the body of published papers.
● Automated extraction using large language models introduces a risk of misattribution of variant-cancer-treatment relationships, and without systematic manual validation of outputs, the precision of identified associations at scale remains difficult to fully characterise.

## Research in context and introduction

Precision oncology aims to tailor cancer treatments for a tumour’s unique genetic, molecular, or cellular characteristics, thus enabling more effective and targeted therapeutic interventions than conventional one-size-fits-all approaches.^1^ Advances in next-generation sequencing (NGS) have facilitated the detection of genetic alterations, providing approaches for identifying actionable variants that guide diagnostic classification, prognosis, and personalised treatment decisions.^2^ Large-scale studies integrating NGS with clinical data have demonstrated the clinical relevance of various genetic alterations, including missense mutations, structural variants, and functional signatures such as microsatellite instability (MSI) and homologous recombination deficiency (HRD).^3,4^ Numerous clinically relevant alterations have been successfully targeted with precision therapies, such as *BRAF* V600E, an oncogenic variant occurring in approximately 6% of all cancers and 40-50% of melanomas, where it activates the MAPK/ERK pathway and has demonstrated clinical responsiveness to combined BRAF and MEK inhibition.^5^ Similarly, common *EGFR* alterations in non-small cell lung cancer (NSCLC), including exon 19 deletions and the L858R point mutation, are effectively treated with tyrosine kinase inhibitors (TKIs).^6–8^ Alterations in *BRCA*1/2, particularly in prostate, breast, ovarian and pancreatic cancer, have shown responsiveness to PARP inhibitors, offering clinical benefit across these tumour types. ^9,10^ Furthermore, multiple other oncogenic drivers leading to drug approval have been identified in oncology including mutations, fusions and gene amplifications (e.g. MET exon 14, EML4-ALK, HER2, ALK). However, many variants remain poorly understood, especially in rare cancers or underrepresented populations, limiting their clinical utility due to unclear biological and functional relevance.^11^

In addition, therapeutic relevance of variants can vary across tumour types. Factors like pathway dependence, homologue-conferred redundancy, synthetic lethality, co-mutations, resistance mechanisms (e.g., MET amplification), microbial cues, and the tumour microenvironment influence treatment response.^12–15^ The presence of a therapeutically-actionable oncogenic variant alone does not guarantee efficacy, emphasizing the need for tumour-context-aware interpretation.^16^ Clinical utility is often constrained by biological complexity, tissue-specific signalling, pharmacokinetics, and side effects produced by on-target-off-cancer or off-target effects.^17^ For example, *BRAF* V600E responds well to *BRAF* inhibitors in melanoma but requires *EGFR* blockade in colorectal cancer to overcome compensatory signaling.^18^ In addition, *BRAF* V600E targeting is frequently limited by acquired drug resistance, necessitating further-line or salvage therapeutic options accordingly; in melanoma, this has contributed to favouring checkpoint-based immunotherapy in the first-line setting for many patients. Although activating EGFR mutations are established therapeutic targets in NSCLC, the clinical efficacy of EGFR-targeted therapies in colorectal cancer is limited by downstream pathway activation, most commonly through KRAS, NRAS, or BRAF alterations, which can drive resistance independent of EGFR signalling.^19^ Therapeutic tractability also depends on the physiological function(s) of the affected gene(s): oncogenes (e.g., *EGFR, BRAF, ALK, MET*) are generally more targetable, while tumour suppressor genes, typically disrupted by loss-of-function mutations or loss-of-heterozygosity, remain challenging to address and often are indirectly targeted based on secondary vulnerabilities such as DNA repair deficiency or a hypermutation phenotype.^20^ *TP53*, the most frequently reported gene in our dataset (70,093 articles) and the gene with the second-highest number of distinct variants (409), exemplifies this: it ranked only eleventh among genes with verified variant–treatment–cancer associations, which were dominated by strong tumour drivers such as *EGFR*, *BRAF* and *KRAS*, reflecting that TP53 mutations, as is typical of tumour suppressors, are spread across many positions rather than concentrated in actionable hotspots. Some alterations are not targeted directly but create vulnerabilities that can be exploited therapeutically, for instance, *BRCA1/2* mutations predict benefit from platinum-based chemotherapy and PARP inhibitors through impaired homologous recombination repair^21^, while high microsatellite instability or mismatch repair deficiency (MSI-H/dMMR) and high tumour mutational burden (TMB) predict response to immune checkpoint blockade in tumour-agnostic settings.^22,23^ Significant gaps persist, especially for understudied variants such as atypical and uncommon mutations. A key example is Cancer of Unknown Primary (CUP), a diagnostically and therapeutically challenging entity with poor prognosis. Molecular tumour profiling, including DNA methylation analysis, is often used in an attempt to identify cell or tissue origin, however, identification of a presumed primary tumour does not necessarily translate into improved clinical outcomes.^24^ A less recognised challenge is the impact of clinical testing patterns and timing of molecular data collection. For example, AR alterations in prostate cancer are often reported due to frequent NGS testing in advanced, castration-resistant stages.^25^

Genomic knowledge bases, such as COSMIC^26^, CIViC^27^, ClinVar^28^, and OncoKB^29^, provide curated resources supporting the interpretation of tumour-detected gene variants, helping clinicians assess clinical significance and inform targeted treatment strategies.^30^ Despite growing efforts to curate and standardise variant interpretation, knowledge remains fragmented and inconsistently represented, with annotations spanning Human Genome Variation Society (HGVS), DNA changes, protein changes, and SNP IDs, complicating interpretation across platforms,^31^ and incomplete, due to the high volume of literature, the cost of manual curation, and scalability limitations.^32,33^ As a consequence, clinicians often consult multiple databases and the primary literature in parallel to interpret variants, making the process time-consuming and non-standardised.^34^ In molecular tumour boards (MTBs), clinicians are often confronted with multiple variants of unknown significance/actionability, rare variant-tumour entity contexts, and inconsistent nomenclature across reports, requiring significant time to investigate, potentially delaying decisions and treatment. There is therefore an urgent need for automated approaches to support information retrieval for clinical variant interpretation.

Traditional automated information retrieval methods, including keyword-based searches, named entity recognition (NER) models such as SciSpaCy^35^ and specialised transformer-based approaches such as BERT^36^, have improved biomedical entity extraction.^37^ However, these task-specific methods often struggle with complex variant mentions, ambiguous context, and rare event detection, and tend to perform poorly on unseen datasets.^38,39^ Modern general-purpose large language models (LLMs) have been rapidly changing biomedical text processing in recent years.^40,41^ LLMs provide semantic flexibility and contextual understanding, enabling them to capture nuanced variant expressions and resolve ambiguities that limit traditional approaches.^42^ Despite this progress, most published variant information extraction approaches still rely on machine learning or rule-based systems.

To address this gap, we developed an automated pipeline using LLM techniques to extract, standardise, and contextualise variant information from unstructured biomedical abstracts. *Variantscape* is designed to support structured evidence exploration in molecular oncology and MTBs by organizing and linking literature-derived associations to their underlying abstracts. The pipeline addresses persistent issues such as inconsistent nomenclature, rare variant detection, and database gaps. By incorporating variant-treatment-disease co-occurrence determination and LLM-based verification, context-rich insights for scientific lead generation are gained, allowing clinicians to discover associations, including rare or underreported ones, without time-consuming manual searches.

## Methods

All analyses (Figure 1) were conducted using Python 3.11.5. The complete code is available on a GitHub repository^46^, while the datasets are publicly available on Zenodo.^47^

**Figure 1:**
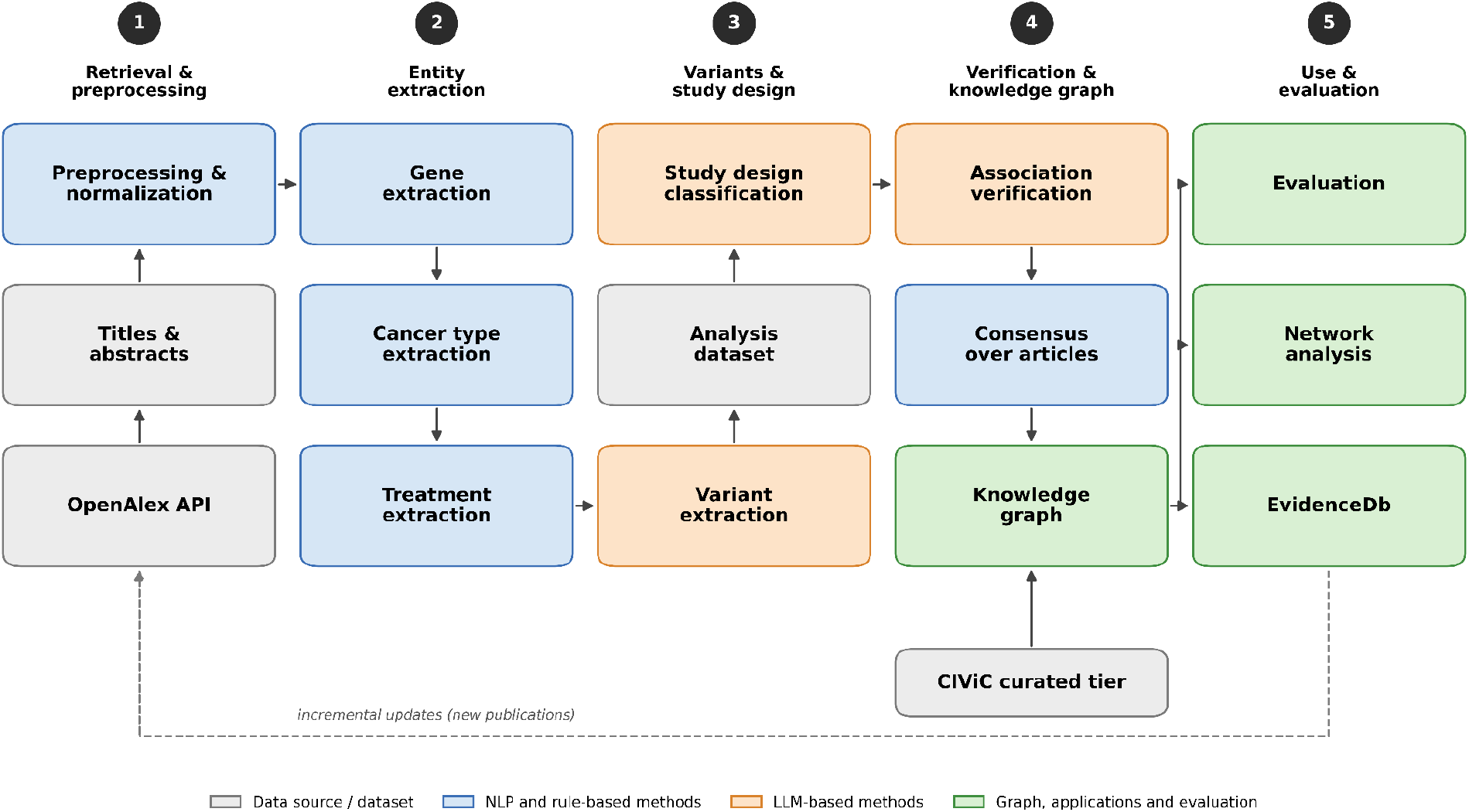
Workflow of computational NLP- and LLM-based extraction of biomedical entities, i.e., cancer types, treatments, study types and molecular variants, and analysis pipeline.

### Literature retrieval

Biomedical publications related to cancer research were retrieved from the OpenAlex database using its public API.^48^ A structured, broad search query, “cancer” OR “tumor”/“tumour” OR “carcinoma”, was applied to search titles and abstracts and identify relevant articles published in English between 01/01/2014 and 18/10/2026, on which date the search was executed. Paper titles, abstracts, and additional metadata were extracted, including publication year, authors, referenced citations, and associated research concepts.

### Dataset cleaning and preprocessing

The dataset was cleaned and normalised for quality and consistency. Duplicates were removed using unique identifiers or title-author combinations. Articles missing titles, abstracts, or metadata, as well as non-English content, were excluded. Supplementary materials, corrections, tables, figures, and artifacts were filtered using keyword and regex patterns. Texts that were unusually short or long were excluded, along with withdrawn articles and non-original content. HTML artifacts were removed, and titles and abstracts were standardised. Publication trends were analysed temporally.

### Information extraction

#### Transformer-based NER for gene extraction

Publications were selected using all genes in the OncoKB Cancer Gene List^29^ or the CIViC knowledgebase^27^ (1,560 genes, retrieved on 18 October 2026, to cover both established and emerging oncology-relevant genes. After preprocessing, articles mentioning these genes were filtered for focused variant analysis. Rule-based string matching and SciSpaCy^35^ underperformed, missing relevant mentions and generating false positives due to limited contextual understanding.^50^ LLMs were similarly performant but less efficient than a BioBERT-based method.^50^ As a result, the BioBERT model alvaroalon2/biobert_genetic_ner^51^ was used. Gene synonyms were retrieved via the MyGene.info API^52^, including symbols, aliases, and protein associations. Titles and abstracts were processed using overlapping windows of up to 510 tokens (stride 256 tokens). Articles without a match to a gene were excluded.

#### Categorization of entities in articles

Various approaches were evaluated to categorise biomedical entities and categories, including transformer-based NER models and LLMs. Assessed models included BERT^36^, pretrained on biomedical corpora, and SciSpaCy models^35,54^. Model selection was based on validation performance using standard metrics such as precision, recall, accuracy, and F1-score.

##### NER for automated cancer type identification

A multi-step NER pipeline was developed to extract, normalise, and categorise cancer types from unstructured text. Cancer mentions were identified in titles and abstracts using two SciSpaCy models: “en_ner_bionlp13cg_md,” optimised for cancer terms, and “en_ner_bc5cdr_md,” trained on broader disease entities.^54^ A reference list of cancer types and synonyms was created from CIViC, mapped to Disease Ontology ID^55^ and MONDO^56^ terms, and expanded via the MONDO API.^57^ Extracted mentions underwent rule-based cleaning and were matched to canonical terms using direct and fuzzy matching^53^ against a CIViC-MONDO dictionary. Standardised types were linked to ontological parents using Disease Ontology metadata^58^ and normalised to the OncoTree hierarchy.^49^ Articles without matches were excluded.

##### Ontology-guided NER for automated treatment identification

The treatment extraction involved querying the CIViC API^59^ for a curated list of cancer therapies, including names, NCIt identifiers, and aliases. The NCIt API^60^ was used to retrieve parent concepts and treatment categories. A hybrid rule-based method, combining regular expressions and fuzzy matching, identified treatments. Ambiguous or short aliases were filtered, and treatment presence was recorded.

##### LLM-based classifier for automated study type categorization

To classify study types, an LLM-based method was used with DeepSeek-V4.1-Flash through the General Classifier framework and DeepInfra API.^60–63^ Categories from the Ontology of Biomedical Investigations (OBI)^64^ were consolidated into eight final categories, including “clinical study,” “observational/RWE study,” “case report study,” “*in vitro* study”, “*in vivo*/animal study,” “*in silico* study,” “systematic review study,” or “other.”. Prompt engineering was tested on a subset (Supplementary table 3). Abstracts were embedded using SciBERT^65,66^ and projected into two dimensions with UMAP, generating an interactive 2D cluster map with category-coloured points and hover data for titles and publication years.^67^

### LLM-based molecular variant extraction

A methodological innovation of this study was extracting specific gene variants from biomedical text using LLMs, as conventional NLP methods have proven insufficient.^42^ To contextualise the pipeline’s scope and novelty, we compared existing variant tools and databases, including OncoKB^29^, CIViC^27^, ClinVar^28^, COSMIC^26^, VarSome^68^, tmVar^43^, PubTator^44^, GenomeNexus^69^, and PeCanPIE^70^, focusing on extraction capabilities, contextual sensitivity, and scalability, with key distinctions detailed in the Discussion. Variant extraction used DeepSeek-V4.1-Flash via DeepInfra.^49^ The prompt instructed the model to extract HGVS notations, protein changes, and SNP IDs. Extracted variants were standardised using preprocessing steps including prefix removal, amino acid canonicalization, exon mapping, and keyword filtering. Synonym coverage and validation were enhanced through APIs from CIViC, ClinVar, and Ensembl Variant Recoder.^71^ Matched variants were standardised as unique variant-gene pairs. Unmatched variants were retained in original form. For each article, the extracted variants, variant-cancer pairs and variant-cancer-treatment triples were retained.

### Association network, LLM-based verification and ranking

Each candidate co-association among variants, treatments, and cancer types was checked against the article title and abstract by an LLM (DeepSeek-V4.1-Flash). The model was prompted to determine whether the publication explicitly reported the association as a finding of the study in the text. The model also assigned a relation (sensitive, resistant, no response, diagnostic, prognostic, reported without direction, or not reported), and quoted the supporting text verbatim. “Resistant” was restricted to resistance caused or predicted by the variant itself, not resistance arising through other mechanisms in patients carrying it, such as acquired MET amplification during targeted treatment with TKIs.^92^

An undirected network of variants, cancer types and treatments was built using NetworkX^73^, with edges between entities of different types. Articles were weighted by study design using a scale informed by the Oxford Centre for Evidence-Based Medicine (OCEBM) levels of evidence^72^: Level I (systematic review study, weight = 1·0), Level II (clinical study, weight = 1·0), Level III (observational/RWE study, weight = 0·9), Level IV (case report study, weight = 0·9), and Level V (in vivo/animal study = 0·8, in vitro study = 0·7, in silico study = 0·6). The weight of an edge is the sum of the weights of the articles verified to report the association; a verified triple supports its three pairwise edges. Co-mentions that were not verified were kept as a separate, lower-ranked tier. For each association, the relations assigned across publications were combined into an overarching consensus. If all articles agreed, their relation was used. Otherwise, an association was labelled sensitive, or resistant/no response, if at least 60% of the articles reporting a treatment response agreed, and “mixed” if not. The number of articles per relation was retained.

Expert-curated associations from CIViC^27^ (evidence levels A–D and AMP/ASCO/CAP tier I–II assertions) were added with their provenance and a score based on their evidence level. In the online web interface, associations are ranked in tiers: curated associations first (by curated score), then verified literature associations (by edge weight). Co-mention-only associations are shown only on request and are marked as unverified. Literature associations are highlighted if their weight is at least 3 and in the top 20% for the query. Searches for a cancer type include its OncoTree subtypes, and searches for a specific variant include evidence for its hotspot codon (e.g. BRAF V600 for V600E. Each association links to the articles that report it, along with the supporting quotes.

Top co-associations were manually validated by three authors (MB, TP, MF) using DrugBank^74^ ClinicalTrials.gov^75^, PubMed^76^, and Google Scholar. Manual review assessed whether the association was supported by at least one primary source describing the variant–treatment relationship in the given cancer context. Under-characterised variants and treatments were identified via targeted search and literature review. A Flask-based web tool integrates a genomics database (CIViC API^45^) and a literature search, providing an LLM-based synthesis of the findings across these different evidence types for a given query. The tool was publicly deployed on a UWSGI server.

### Evaluation of the association network

The literature-derived associations were evaluated in three ways. First, recall was assessed against two curated reference sets: predictive evidence and assertions from CIViC (evidence levels A–D and AMP/ASCO/CAP tier I–II; snapshot of 18 October 2026), and the Molecular Oncology Almanac (MOAlmanac)^91^ (release 2026-05-07). Each reference association was classified as verified in at least one article (for the same cancer type, or for an OncoTree parent or subtype, or the variant’s hotspot codon), as co-mentioned only, or as not found. For associations found, we assessed whether the consensus relation agreed with the reference (sensitive vs. resistant/no response). Second, ranking quality was assessed with the curated tier switched off. For each variant–cancer pair with a sensitive treatment at a strong evidence level (CIViC level A/B or tier I; MOAlmanac FDA-approved or guideline), we recorded whether a reference treatment was among the top 1, 3 or 6 sensitive treatments listed, and computed the mean reciprocal rank. Third, the precision of novel associations was assessed manually. Verified variant–cancer–treatment associations that matched no reference association were sampled in three strata by number of supporting articles (1, 2–3 and ≥4; 20 per stratum). Two independent annotators [MB and TP] judged, from the supporting title and abstract, whether the association and its relation were reported, and whether the variant, cancer type and treatment were correct, following written guidelines. Precision was reported per stratum with Wilson 95% confidence intervals and overall after re-weighting by stratum size; agreement between annotators was measured with Cohen’s kappa.

## Results

### Performance summary of information extraction pipeline

Each component of the extraction pipeline was evaluated against a manually annotated ground truth. Gene extraction using the BioBERT-based *biobert_genetic_ner* model showed the strongest performance (F1-score: 0·98, Recall: 1·00).^50^ Cancer type extraction with SciSpaCy models and ontology-guided normalization yielded F1-score: 0·85, Recall: 0·77 (Supplementary Table 1). Treatment extraction via hybrid ontology and fuzzy matching achieved F1-score: 0·95, Recall: 0·95 (Supplementary Table 2). Variant extraction by LLM showed the strongest performance (F1-score: 0·95, Recall: 0·92).^42^ Study type classification reached F1-score: 0·92, Recall: 0·92 (Supplementary Table 3). Variant-cancer-treatment classification using DeepSeek-V4.1-Flash was evaluated against curated knowledge bases, with the literature-derived associations recovering 54% of CIViC and 61% of MOAlmanac associations (82% of CIViC level A evidence and 89% of FDA-approved MOAlmanac associations), with 87–92% agreement on the direction of response; manual annotation of a stratified sample of associations not present in either knowledge base estimated a precision of 0.82–0.86 (Supplementary Table 4).

### Study selection and characteristics

A total of 3,727,471 cancer-related biomedical articles published between 2014 and 2026 were retrieved from the OpenAlex database. Following preprocessing, 829,517 records (22.3%) were excluded due to duplicates (4.9%), missing metadata (1.7%), non-English language (2.1%), withdrawn articles (0.1%), nonsensical titles (0.1%), extreme length (7.0%), non-original research (0.9%), normalization errors (<0.1%), and misclassified supplementary materials (5.5%). The cleaned dataset contained 2,897,954 articles. Temporal analysis showed steady growth from about 160,000 articles in 2014 to over 300,000 by 2025 (Supplementary Figure 1).

### Transformer-based gene extraction

From the pre-processed dataset of 2,897,954 publications, the BioBERT model identified 886,195 articles (30.6%) mentioning genes from the oncology gene set in titles or abstracts. As a result, 2,011,759 articles (69.4%) were excluded. In the filtered set, *TP53* was most frequently mentioned (70,093 articles; 7.9%), followed by *ERBB2* (HER2; 67,349; 7.6%), CD274 (PD-L1; 59,409; 6.7%), *EGFR* (55,717; 6.3%), *AKT1* (52,800; 6.0%), *PDCD1* (PD-1; 49,464; 5.6%)*, PIK3CA* (49,414; 5.6%) and *IL6* (46,634; 5.3%), with some bias from pathway-focused and immunotherapy research, as protein-level biomarkers such as PD-L1 and HER2 are reported under their gene (Figure 2A). Most articles (608,008; 68.6%) referenced one (395,364; 44.6%) or two genes (212,644; 24.0%), with an average of 2.36 genes per article; 159,988 articles (18.1%) mentioned four or more genes (Supplementary Figure 2). Most articles (608,008; 68.6%) referenced one (395,364; 44.6%) or two genes (212,644; 24.0%), with an average of 2.36 genes per article; 159,988 articles (18.1%) mentioned four or more genes (Supplementary Figure 2).

**Figure 2:**
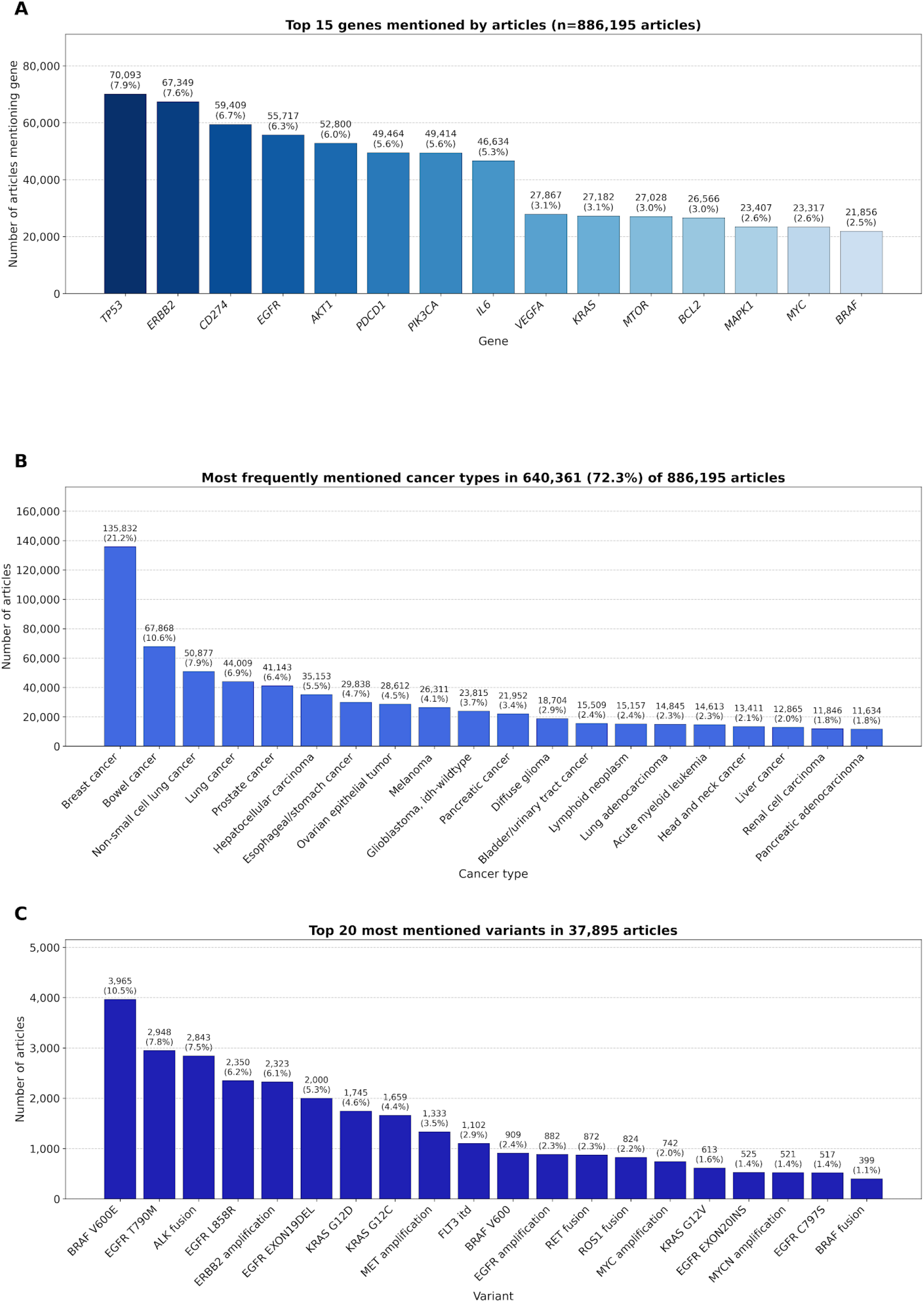
A) Top 15 most frequently mentioned genes across 886,195 biomedical cancer-related publications mentioning genes of the oncology gene set. B) Distribution of the most frequently mentioned cancer types identified in 640,361 articles (72.3% of the 886,195 publications). C) Top 20 most frequent variants in the analysis dataset of 37,895 articles.

### Categorization of biomedical entities

NER-based cancer extraction identified at least one cancer type in 640,361 (72.3%) of the 886,195 screened articles. A total of 210 distinct types were found, with the most frequent being breast cancer (135,832; 21.2%), bowel cancer (50,877; 7.9%), lung cancer (44,009; 6.9%), prostate cancer (41,143; 6.4%), hepatocellular carcinoma (35,153; 5.5%), oesophageal/stomach cancer (29,838; 4.7%), ovarian epithelial tumour (28,612; 4.5%) and melanoma (26,311; 4.1%), reflecting incidence and overall research activity, including prominence in targeted or immunotherapy development (Figure 2B).

Of the 37,895 articles in the final analysis dataset, most were classified as clinical studies (9,177; 24.22%), followed by in vivo/animal studies (6,598; 17.41%), in vitro studies (6,397; 16.88%), observational/RWE studies (6,101; 16.10%), case reports (5,620; 14.83%), systematic reviews (2,622; 6.92%) and in silico studies (724; 1.91%). A small subset (649; 1.71%) was labelled “undefined” due to ambiguity or unmatched design types (Figure 3, Supplementary Figure 3).

**Figure 3:**
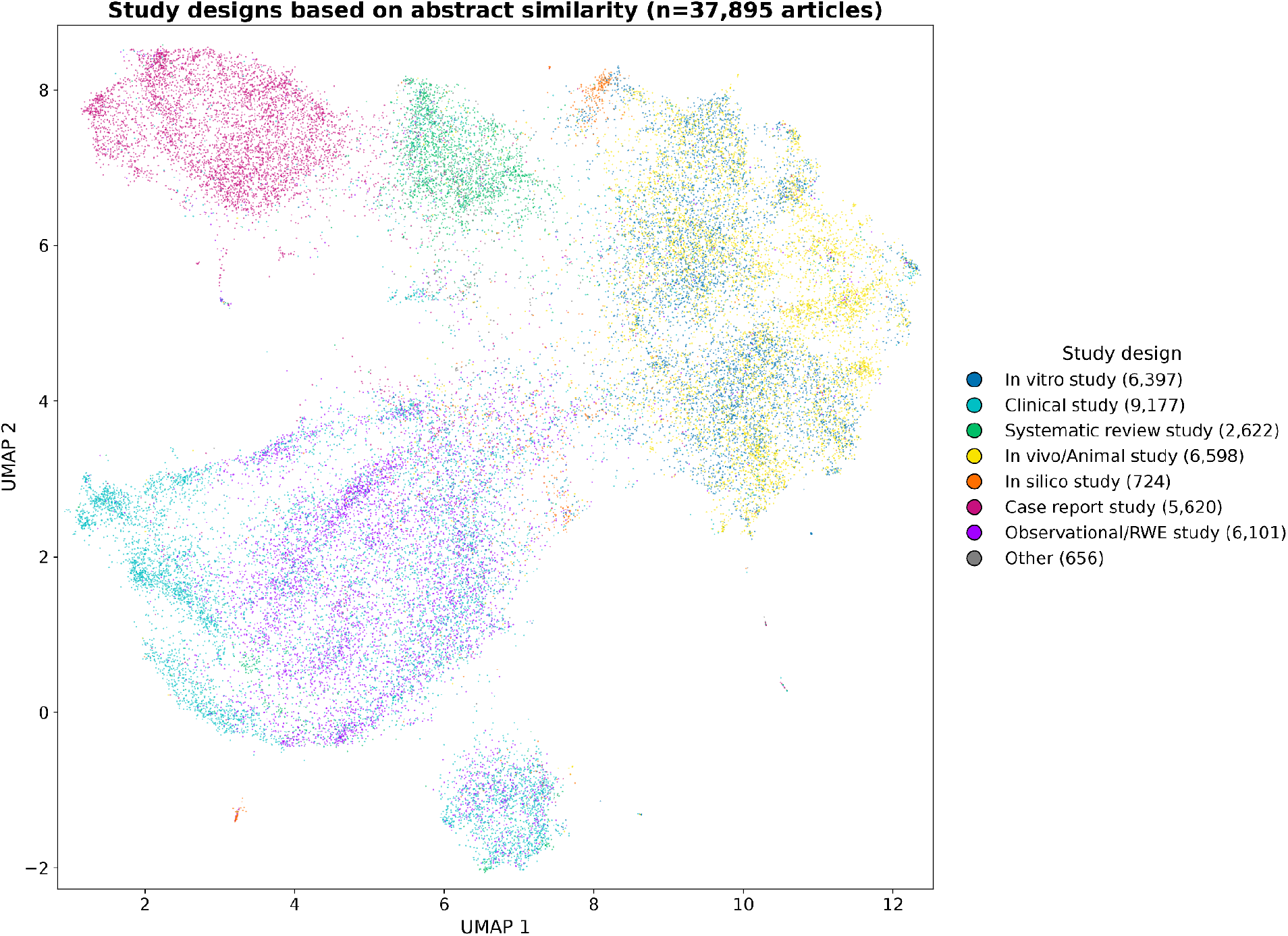
UMAP-based visualisation of embeddings from 37,895 articles by study design category, classified by DeepSeek-V4.1-Flash.

Out of 886,195 gene-containing articles, 365,742 (41.27%) mentioned treatments, while 520,453 (58.73%) lacked mapped references. Besides general treatment classes such as chemotherapy (115,002; 13.0%) and immunotherapy (78,483; 8.9%), the top treatments included cisplatin (19,163; 2.2%), trastuzumab (14,562; 1.6%), paclitaxel (12,987; 1.5%), pembrolizumab (11,928; 1.3%), doxorubicin (11,866; 1.3%) and sirolimus (9,621; 1.1%) (Supplementary Figure 4).

### LLM-based variant extraction

The LLM-based variant extraction pipeline processed 292,287 cancer-related articles that mentioned a gene, a cancer type and a treatment, using DeepSeek-V4.1-Flash. Variants were identified in 37,895 articles (12.96%), while 254,392 (87.04%) lacked variant mentions. This subset represented about 1% of all articles initially retrieved from OpenAlex. After normalisation, 72,435 variant mentions were found, with 11,985 unique variants, indicating that many articles referenced multiple variants. Of the 133,647 variants extracted by the LLM, 54,383 (40.7%) were already in canonical form and 26,512 (19.8%) were normalised (e.g. three-letter to one-letter amino acid notation, fusion and gene alias naming); 44,167 (33.0%) were removed as non-specific terms (e.g. “mutation”, “wild-type”) and 8,585 (6.4%) because they did not occur in the title or abstract. The most frequently mentioned alterations were *BRAF* V600E (3,965; 10.46%), *EGFR* T790M (2,948; 7.78%), *ALK* fusion (2,843; 7.50%), *EGFR* L858R (2,350; 6.20%), *ERBB2* amplification (2,323; 6.13%), *EGFR* exon 19 deletion (2,000; 5.28%), *KRAS* G12D (1,745; 4.60%) and *KRAS* G12C (1,659; 4.38%) (Figure 2C). Most variant mentions (66,865; 92.31%) were linked to genes of the oncology gene set. Of its 1,560 genes, 915 (58.65%) had at least one variant. The remaining 5,570 variant mentions (7.69%) mapped to genes not covered in the oncology gene set (Supplementary Figure 5).

### Co-occurrence analysis

Of the 886,195 articles mentioning a gene of the oncology gene set, 640,361 (72.3%) mentioned at least one cancer type and 365,742 (41.3%) a treatment; 292,287 (33.0%) mentioned all three and were screened for variants. 37,895 (4.3%) mentioned at least one variant in the title or abstract and formed the analysis dataset (Supplementary Figure 6). This dataset included 547 unique treatments, 197 cancer types and 11,985 variants. Besides general treatment classes (chemotherapy, 30.0% of articles; tyrosine kinase inhibitors, 18.6%), the most frequently mentioned treatments were osimertinib (2,526; 6.67%), crizotinib (1,989; 5.25%), trastuzumab (1,500; 3.96%), gefitinib (1,409; 3.72%) and erlotinib (1,381; 3.64%) (Supplementary Figure 7). The most frequent cancer types were non-small cell lung cancer (9,962; 26.29%), lung cancer (4,652; 12.28%), breast cancer (4,469; 11.79%), bowel cancer (3,778; 9.97%) and melanoma (2,434; 6.42%) (Supplementary Figure 8), and the most frequent variants *BRAF* V600E (3,965; 10.46%), *EGFR* T790M (2,948; 7.78%), *ALK* fusions (2,843; 7.50%), *EGFR* L858R (2,350; 6.20%) and *ERBB2* amplification (2,323; 6.13%) (Figure 2C; Supplementary Figure 9).

For each article, all combinations of its variants, cancer types and treatments were verified against the abstract by the LLM (368,042 candidate associations). In total, 170,283 (46.3%) were confirmed as reported by the article with a verbatim supporting quote; 1,321 were rejected because the quoted text was not found in the abstract. 35,224 articles (93.0%) reported at least one verified association, and 26,089 (68.8%) at least one variant–cancer–treatment association. Among the 95,584 verified variant–cancer–treatment associations, 39.7% were labelled sensitive, 16.8% resistant, 2.5% no response and 1.4% prognostic; 39.6% reported the association without a direction. Combined across articles, these yielded 32,292 distinct verified variant–cancer–treatment triples and 17,242 variant–cancer pairs. Aggregating the labels of all articles for 21,177 variant–treatment pairs resulted in a consensus of sensitive for 7,348 pairs (34.7%), resistant for 4,021 (19.0%), no response for 760 (3.6%) and prognostic for 745 (3.5%). A further 7,846 pairs (37.0%) were reported without a direction and 457 (2.2%) were mixed, i.e. the articles disagreed without a 60% majority. The resulting knowledge graph contains 12,729 literature-derived nodes and 95,224 edges, of which 44,644 are supported by at least one verified abstract and the remainder by co-occurrence only (Figure 4). Of the 1,911 curated CIViC associations added as a separate tier, 52.6% were also supported by verified literature. The literature-derived and CIViC classifications agreed in 90.2% of cases where both provided a directional association.

**Figure 4:**
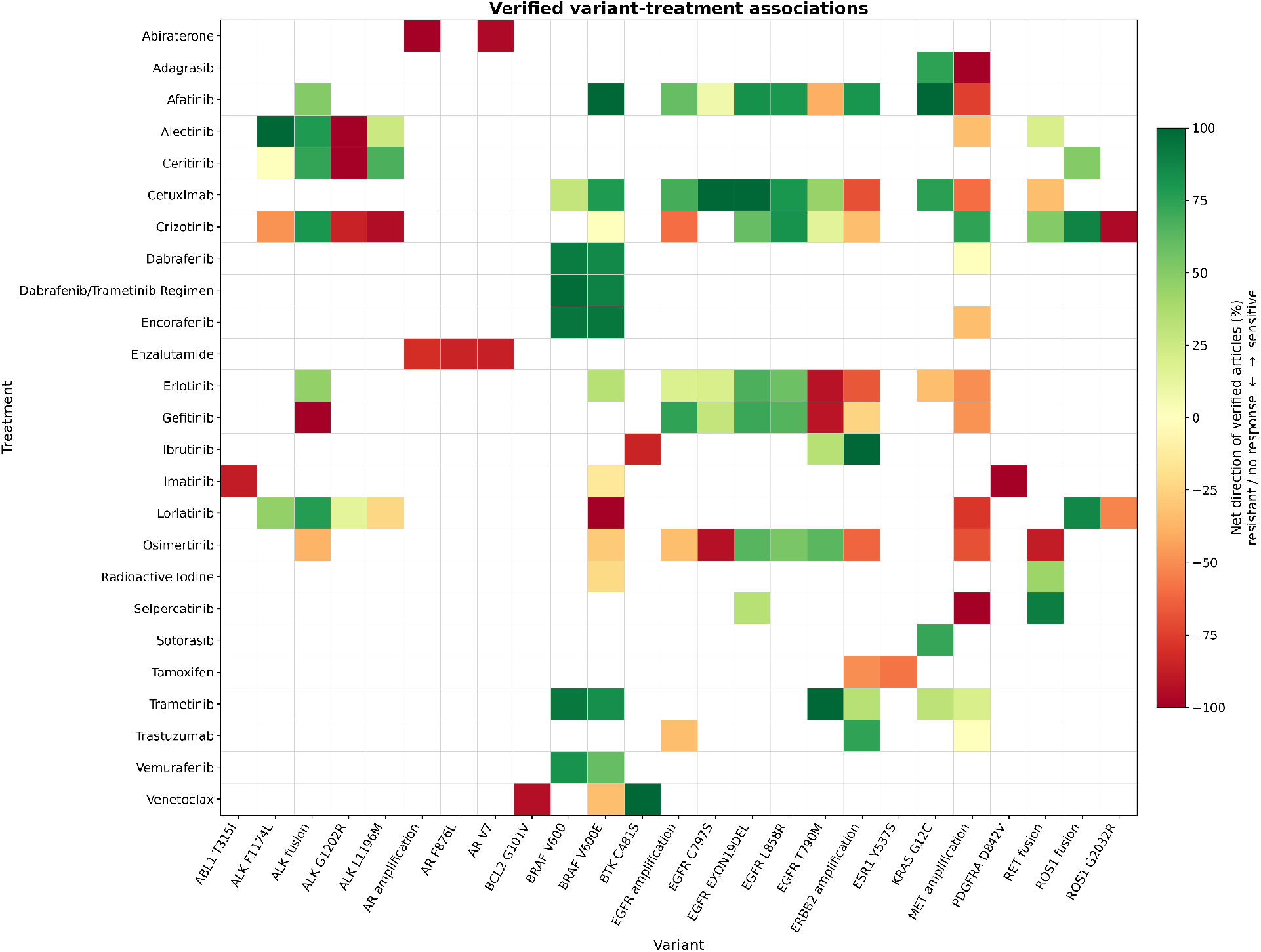
Heatmap of verified variant–treatment associations. The 25 sensitive and 25 resistant associations supported by most articles were selected; colours show the net direction of the verified articles for each pair with at least three such articles (green: sensitive; red: resistant or no response).

Well-established predictive associations were recovered with strong literature support. Beyond the most frequent cancer types, the strongest variant–cancer associations included *BRAF* V600E in anaplastic and papillary thyroid cancer (68.7% and 51.1% of articles), *MYCN* amplification in neuroblastoma (60.9%), *KRAS* G12D in pancreatic adenocarcinoma (56.8%) and *MYC* amplification in medulloblastoma (50.9%). In melanoma, *BRAF* V600E was associated with sensitivity to vemurafenib (463 articles; 288 sensitive), dabrafenib (228) and trametinib (171); in bowel cancer, with sensitivity to encorafenib (197 articles; 149 sensitive) and cetuximab (252). *KRAS* G12C was associated with sensitivity to sotorasib (405 articles) and adagrasib (255) in non-small cell lung cancer. Resistance mechanisms were captured as well: *EGFR* T790M with resistance to gefitinib (314 articles; 214 resistant) and erlotinib (309), but sensitivity to osimertinib (1,014); *EGFR* C797S with resistance to osimertinib (268; 213 resistant); *ALK* G1202R with resistance to crizotinib, alectinib and ceritinib but sensitivity to lorlatinib; and the *AR* splice variant V7 with resistance to enzalutamide in prostate cancer (234). Mixed associations highlighted conflicting evidence, for instance *ESR1* Y537S and D538G with fulvestrant in breast cancer (85 and 56 articles), where sensitivity and resistance were reported in similar proportions.

The variant spectrum differed between cancer types (Figure 5): in non-small cell lung cancer, *EGFR* T790M (22.4% of articles), ALK fusions (18.5%) and *EGFR* L858R (17.5%) dominated; in bowel cancer, *BRAF* V600E (25.3%) and *KRAS* G12C (8.5%); in melanoma, *BRAF* V600E (50.7%); in breast cancer, *ERBB2* amplification (21.5%) and *ESR1* Y537S (7.2%); in prostate cancer, the *AR* splice variant V7 (27.4%) and *AR* amplification (16.6%); and in acute myeloid leukaemia, *FLT3* internal tandem duplications (45.5%).

**Figure 5:**
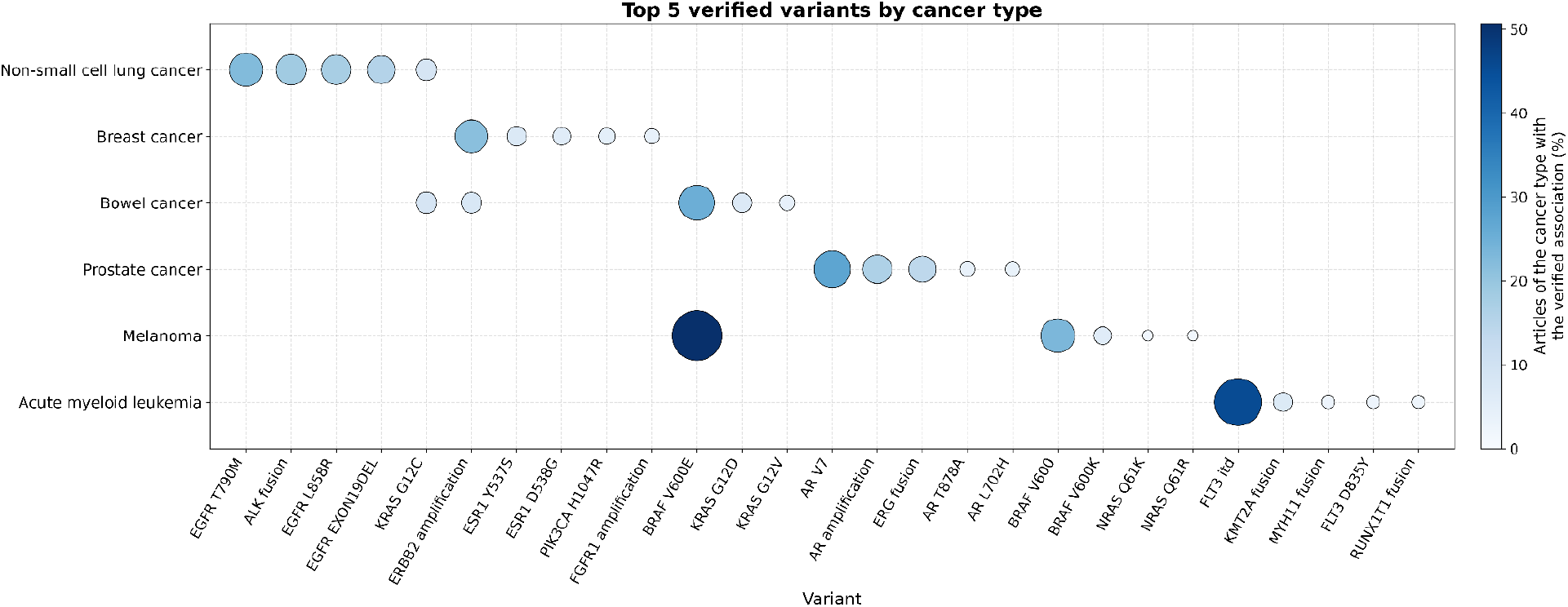
Top five variants by cancer type, by the share of the articles of each cancer type in which the variant–cancer association was verified.

Verified treatment–cancer associations similarly reflected established standards of care, such as imatinib in gastrointestinal stromal tumours and Philadelphia-positive leukaemias, BRAF/MEK inhibitors in anaplastic thyroid cancer and melanoma, and androgen receptor pathway inhibitors in prostate cancer (Supplementary Figure 10).

### Network analysis

The literature-derived network included 12,729 nodes (11,985 variants, 547 treatments and 197 cancer types) and 95,224 edges, of which 44,644 edges, connecting 10,571 nodes, were supported by at least one verified abstract; the remainder reflected co-occurrence only. Adding the curated CIViC tier extended the graph to 12,955 nodes and 96,227 edges. Edges are weighted by the number of articles verified to report the association, weighted by study design. The most connected variants were *BRAF* V600E (304 verified links to 82 cancer types and 222 treatments), *ALK* fusions, *EGFR* T790M, *EGFR* L858R and *EGFR* exon 19 deletions (Table 1). For non-small cell lung cancer (NSCLC), the strongest variant links were *EGFR* T790M (2,268 articles), *ALK* fusions (1,865), *EGFR* L858R (1,763) and *EGFR* exon 19 deletions (1,548).

**Table 1:** Top five “sensitive” and “resistant” treatment associations for EGFR L858R and EGFR T790M in non-small cell lung cancer (NSCLC), and the other cancer types associated with these variants, based on network analysis.

| **Cancer type: Non-small cell lung cancer (NSCLC)** |  |
|---|---|
| **Variant: L858R *EGFR*** | **Variant: T790M *EGFR*** |
| ***Top 5 associated “sensitive” treatments*** *(weighted evidence score)\** |  |
| **Osimertinib: 464.8** | **Osimertinib: 887.5** |
| **Gefitinib: 225.2** | **Rociletinib: 59.0** |
| **Erlotinib: 211.3** | Lazertinib: 29.0 |
| **Afatinib: 153.9** | Aumolertinib: 25.4 |
| **Pemetrexed: 54.7** | Naquotinib: 13.9 |
| ***Top 5 associated “resistant” treatments*** *(weighted evidence score)\** |  |
| Vinorelbine: 7.7† | **Erlotinib: 263.3** |
| Brigatinib: 4.3 | **Gefitinib: 262.1** |
| Atezolizumab: 3.9† | **Afatinib: 163.7** |
| Durvalumab: 3.7† | Icotinib: 20.1 |
| Carboplatin/paclitaxel: 2.8† | Dacomitinib: 19.8 |
| ***Other (non-lung) top 5 cancers associated with the respective variant*** *(weighted evidence score)\** |  |
| **Bowel cancer: 2.3** | Bowel cancer: 3.2 |
| Breast cancer: 1.8 | Glioblastoma, IDH-wildtype: 2.5 |
| Hepatocellular carcinoma: 1.8 | Hepatocellular carcinoma: 2.4 |
| Ovarian epithelial tumor: 1.8 | Pancreatic cancer: 1.8 |
| Cancer of unknown primary: 1.8 | Breast cancer: 1.8 |

**\*Weighted evidence score: number of articles in which the association was verified, weighted by study design. Bold: high-confidence associations (≥ 99th percentile of all verified variant–cancer–treatment associations, score ≥ 29.2). Relations are the consensus over the articles (≥ 60% majority). †"No response" rather than "resistant". General treatment classes (e.g. tyrosine kinase inhibitors, chemotherapy) are not listed.**

For *EGFR* L858R and NSCLC, the strongest treatment links all had a sensitive consensus classification and ranked above the 99th percentile of verified associations, including the TKIs osimertinib (weighted evidence score 464.8; 528 articles), gefitinib (225.2), erlotinib (211.3) and afatinib (153.9) (Table 1). No high-confidence resistance associations were found for *EGFR* L858R. The few resistance or non-response associations (e.g. to atezolizumab and durvalumab) were supported by fewer than ten articles each and related to treatment modalities not specifically targeting *EGFR*-mutant disease. By contrast, *EGFR* T790M had a high-confidence sensitive link with the third-generation TKI osimertinib (887.5; 1,014 articles) and strong resistance links with the first- and second-generation TKIs erlotinib (263.3), gefitinib (262.1) and afatinib (163.7), in line with its role as the main on-target mechanism of acquired resistance to these drugs. Both variants were almost exclusively linked to lung cancers (NSCLC, lung adenocarcinoma and lung cancer); links to other cancer types, such as bowel cancer (*EGFR* L858R: 3 articles; *EGFR* T790M: 4), were rare.

### Rare variant identification and evaluation of treatments

Variant mentions were highly skewed: 30 variants mentioned in more than 300 articles each (e.g. BRAF V600E, EGFR T790M) accounted for 45% of all mentions, while 8,779 of the 11,985 variants (73.2%) were mentioned in only one article and 11,071 (92.4%) in at most five. To assess whether the literature-derived graph adds knowledge for rare variants beyond curated resources, we systematically selected associations between rare variants (≤300 articles) and specific treatments in lung cancers that were verified in at least five articles with a consistent direction (≥60% majority criterion). Of the 109 such associations for individually named variants, 54 (50%) had no predictive evidence for the same variant and drug in CIViC, and a further 17 (16%) were present only as unreviewed submissions. Table 2 shows four examples, whose supporting articles were manually checked. *ROS1* G2032R, the most frequent solvent-front resistance mutation,^78–81^ was associated with resistance to crizotinib (44 of 49 articles), entrectinib (20 of 22; absent from CIViC) and lorlatinib (12 of 19), contrary to the only accepted CIViC record for lorlatinib, but with sensitivity to the next-generation TKIs repotrectinib (21 of 29) and taletrectinib (17 of 18). *EGFR* G724S, an acquired resistance mutation to osimertinib^82–83^ (7 of 9 articles), remained sensitive to afatinib in all seven articles reporting on it, a treatment option not captured in CIViC. *KRAS* Y96D conferred resistance to the *KRAS* G12C-specific inhibitors sotorasib and adagrasib,^84^ and *ROS1* L2086F to all type I *ROS1* inhibitors, while remaining sensitive to the type II inhibitor cabozantinib^85^ (7 of 7 articles). The supporting evidence was predominantly preclinical or from case reports, as expected for rare variants; such associations may inform molecular tumour board discussions and off-label treatment decisions when standard options are exhausted, but require critical appraisal of the underlying studies. All four variants were almost exclusively reported in lung cancer.

**Table 2:**
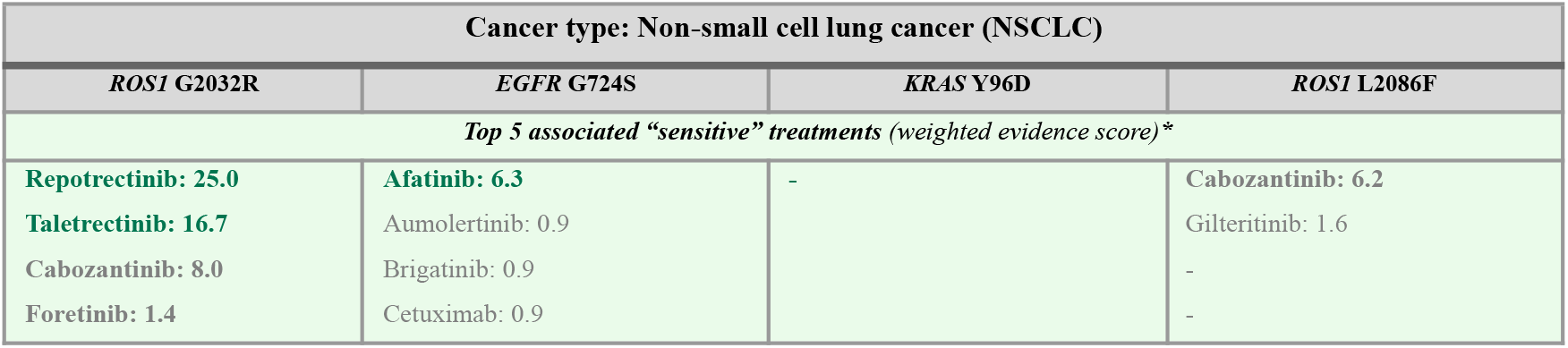
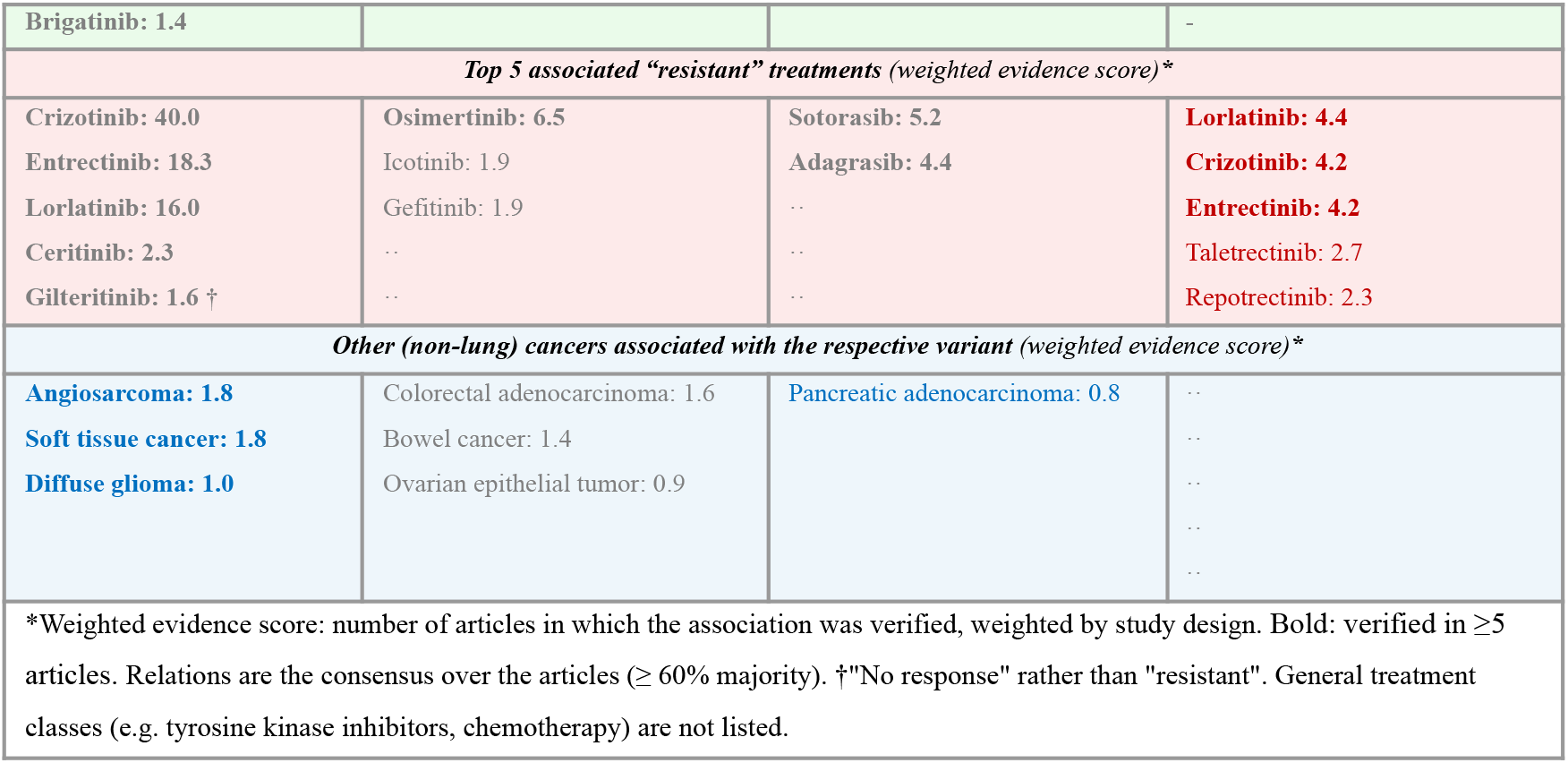
Top five ranked treatment co-associations for rare variants identified in non-small cell lung cancer (NSCLC), along with additional cancer types where these variants might be observed.

| **Cancer type: Non-small cell lung cancer (NSCLC)** |  |  |  |
|---|---|---|---|
| ***ROS1* G2032R** | ***EGFR* G724S** | ***KRAS* Y96D** | ***ROS1* L2086F** |
| ***Top 5 associated “sensitive” treatments*** *(weighted evidence score)\** |  |  |  |
| **Repotrectinib: 25.0** | **Afatinib: 6.3** | - | **Cabozantinib: 6.2** |
| **Taletrectinib: 16.7** | Aumolertinib: 0.9 |  | Gilteritinib: 1.6 |
| **Cabozantinib: 8.0** | Brigatinib: 0.9 |  | - |
| **Foretinib: 1.4** | Cetuximab: 0.9 |  | - |
| **Brigatinib: 1.4** |  |  | - |
| ***Top 5 associated “resistant” treatments*** *(weighted evidence score)\** |  |  |  |
| **Crizotinib: 40.0** | **Osimertinib: 6.5** | **Sotorasib: 5.2** | **Lorlatinib: 4.4** |
| **Entrectinib: 18.3** | Icotinib: 1.9 | **Adagrasib: 4.4** | **Crizotinib: 4.2** |
| **Lorlatinib: 16.0** | Gefitinib: 1.9 | .. | **Entrectinib: 4.2** |
| **Ceritinib: 2.3** | .. | .. | Taletrectinib: 2.7 |
| **Gilteritinib: 1.6 †** | .. | .. | Repotrectinib: 2.3 |
| ***Other (non-lung) cancers associated with the respective variant*** *(weighted evidence score)\** |  |  |  |
| **Angiosarcoma: 1.8** | Colorectal adenocarcinoma: 1.6 | Pancreatic adenocarcinoma: 0.8 | .. |
| **Soft tissue cancer: 1.8** | Bowel cancer: 1.4 |  | .. |
| **Diffuse glioma: 1.0** | Ovarian epithelial tumor: 0.9 |  | .. |
|  |  |  | .. |
|  |  |  | .. |

\*Weighted evidence score: number of articles in which the association was verified, weighted by study design. Bold: verified in ≥5 articles. Relations are the consensus over the articles (≥ 60% majority). †"No response" rather than "resistant". General treatment classes (e.g. tyrosine kinase inhibitors, chemotherapy) are not listed.

### Interactive web tool for querying variant-cancer-treatment co-associations

To translate our findings into a usable resource, we developed the publicly accessible *Variantscape* web tool for interactive exploration of the network analysis and variant, treatment, and cancer associations identified in this study. Users can search by variant, gene, or cancer type to retrieve co-associated treatments and other linked cancers. This interface supports exploratory navigation of literature-derived associations and may aid hypothesis generation in research. The tool is freely available at https://evidencedb.hastingslab.org/variantscape^86^.

Potential applications include the following:

● Initial screening and triage: Rapidly identifying candidate therapies or clinical trials co-mentioned alongside a rare variant in a specific tumour type.
● Evidence surfacing: Retrieving the most relevant literature supporting a given variant–cancer–treatment association.
● Gap identification: Highlighting variant–cancer pairs with sparse, emerging, or conflicting literature signals.
● Harmonisation: Standardising heterogeneous nomenclature (HGVS, protein changes, SNP IDs) to facilitate structured discussion.
● Curation support: Detecting emerging literature signals that may warrant expert review or inclusion in curated knowledge bases.
● Standardising evidence tier allocation of treatment recommendations: Leveraging semi-automated evidence tier classification of particular alteration-drug matches based on the level-of-evidence of supporting data in alignment with established classification systems such as ESMO-ESCAT^81^and the German NCT^82^.

Primary users include translational researchers, interdisciplinary MTB personnel, and curation teams who require rapid evidence discovery and contextual overview rather than automated treatment recommendations.

## Discussion

In rare and understudied cancers, robust prospective trial evidence is often limited or infeasible to generate, and curated databases may not keep pace with emerging evidence from the literature. As precision oncology increasingly uses molecular stratification, predictive biomarkers, and individualised treatments, traditional large-scale trials may become less important for therapeutic development, while the significance of real-world evidence studies is rising correspondingly. To help support targeted evidence retrieval and candidate ranking, we developed an LLM-supported method to extract potential variant-treatment-cancer co-associations across tumour types, aiding hypothesis generation where direct evidence is lacking. Our approach identified many plausible treatment-variant, variant-cancer, and treatment-cancer co-associations. Recall of known associations was highest for well-established evidence (e.g. 96% of CIViC tier I-A assertions and 89% of FDA-approved associations in MOAlmanac) and declined in less-established evidence. Although each association in the graph was verified as reported in an abstract, being reported does not establish clinical validity: abstracts summarise heterogeneous study populations and designs, rare-variant associations rest mostly on preclinical studies and case reports, and verification is limited to titles and abstracts. Variant frequencies in literature may also reflect sampling and testing biases rather than true biological prevalence. This extends to other tumour types as well, even though broad NGS testing at diagnosis is increasing to (i) expand treatment options, (ii) inform first-line treatment, and (iii) enable planning of subsequent therapies. Clinical studies, especially randomised-controlled trials, often include standard-of-care comparators, which can inflate mention frequencies regardless of actual treatment efficacy. In addition, the number of articles supporting an association reflects research activity and publication bias as well as the strength of the evidence, which the study-design weighting only partly corrects. Where the literature disagrees, associations are flagged as mixed rather than resolved. Clinical validation remains essential for assessing relevance to the individual patient.

Our study is situated in the context of other approaches to extract variant mentions from published texts. As an example, tmVar^43^ used a machine learning approach to detect sequence variants in PubMed abstracts based on HGVS nomenclature, but it primarily captures isolated mentions without broader context. PubTator 3.0^44^ expanded biomedical entity extraction across over a billion PubMed annotations, but does not apply LLMs directly to unstructured text, precluding detection of ambiguous or novel variants. VarSomes^45^ improved retrieval of variant-related publications, but it relies on pre-annotated datasets and lacks dynamic extraction capabilities. More recently, fine-tuned language models with manual annotation have improved variant recognition, though most focus on surface-level labelling rather than deeper interpretation or relationship extraction.^26^ Our study addresses this gap, enabling scalable, context-aware extraction of heterogeneous and novel variant mentions. Unlike manually curated resources (Supplementary Table 5), *Variantscape* covers the full abstract-level literature and updates automatically, at the trade-off of lower precision than expert curation.

Nevertheless, our study has several noteworthy limitations. First, we analysed only titles and abstracts, as full-text access is often restricted. Thus, variants mentioned solely in full texts, including tables and supplementary materials, were not captured. Still, abstracts offer a consistent, scalable foundation for mining and discovery. Second, we only processed publications from 2014 to 2026. There is a trade-off to navigate here: the risk of missing earlier-reported associations must be balanced against a risk that some associations may be outdated due to rapid diagnostic and therapeutic advances and changes in clinical practice and guidelines. Moreover, the fully automated pipeline enables updates with new data arising from broad NGS testing of tumour tissue and liquid biopsies, MTB panels, and innovative targeted drugs becoming more established. Third, publication bias affects our results; well-studied genes, cancer entities and therapies dominate, while rare variants or underrepresented populations are less frequently reported. Fourth, the mention of particular variants in titles and abstract does not necessarily imply that these variants were subject to investigation in these papers; indeed, they may only have been mentioned as part of the study’s eligibility criteria or within the context of molecular disease stratification/patient subgrouping. Fifth, compound mutations and treatment combinations pose particular challenges to *Variantscape*, which retrieves one mutation/one treatment at a time, thus possibly withholding critical context-dependent molecular information. Finally, it is important to emphasise that the pipeline only extracts associations that should be viewed as preliminary and require expert human validation before informing clinical decisions.

Our approach has potential for further integration with additional data types and analytical tools. Future work will include full-text mining, at least to the extent that these texts are available, to capture deeper variant annotations, structured therapeutic evidence, and contextual insights not available in titles and abstracts. Expanding extraction to molecular tumour signatures (e.g., MMR, MSI, TMB), and compound biomarkers would improve biological and therapeutic scope. Integrating literature-derived associations with curated databases and real-world datasets could support stronger hypothesis testing and validation. Linking these findings to clinical outcomes may also help prioritise leads for translational research, clinical follow-up, and quality control/monitoring. Finally, progress will also depend on cultural shifts in scientific publishing, including more structured variant reporting, open-access availability, and improved metadata. These changes will be essential to ensure that promising findings can more rapidly and efficiently lead to clinically meaningful insights.

## Conclusion

This study demonstrates the feasibility and value of using LLMs and other NLP methods in combination to systematically extract and explore variant, treatment, and cancer co-associations from biomedical literature. Automation is essential for scaling extraction, maintaining accuracy, and keeping pace with the rapid growth of scientific knowledge. Leveraging advanced machine learning, our pipeline identified thousands of meaningful associations, including canonical and rare variants often underrepresented or hard to retrieve manually or via standard databases. Although limited to titles and abstracts, the method provides a scalable foundation for literature-based variant discovery and maps the evolving molecular oncology landscape.

The interactive, open-access web tool translates these findings into a resource supporting hypothesis generation, early exploration of literature signals, and potential therapeutic applications, especially for rare variants. Despite limitations and need for clinical validation, this approach offers a practical start for enhancing variant interpretation and closing gaps in curated knowledge. As biomedical literature and molecular tumour profiling expand, automated tools like *Variantscape* will be critical for turning unstructured evidence into structured insights reflecting variant complexity and advancing precision oncology.

## Data Availability

The datasets generated and/or analysed during the current study are available in a Zenodo repository and can be accessed via this link: https://zenodo.org/records/15268056.
The underlying code and validation datasets for this study is available in a GitHub repository and can be accessed via this link: https://github.com/hastingslab-org/variantscape.

https://github.com/hastingslab-org/variantscape

## Author contributions

Conception and design: MW, JH.

Data collection, analysis, and statistics: MW, ABS, TN, JH.

Data interpretation: MW, ABS, MB, TP, MF, CR, JH.

Wrote the first draft of the paper: MW.

Wrote the final version of the paper: MW, ABS, MB, TP, TN, MF, CR, JH.

Approved the paper for submission and publication: MW, ABS, MB, TP, TN, MF, CR, JH.

## Data Sharing Statement

The datasets generated and/or analysed during the current study are available in a Zenodo repository and can be accessed via this link: https://zenodo.org/records/15268056.

The underlying code and validation datasets for this study is available in a GitHub repository and can be accessed via this link: https://github.com/hastingslab-org/variantscape.

## Research Ethics Approval: Human Participants

The study uses pre-existing publicly available data and does not collect data from humans or animals.

## Patient and Public Involvement

Patients or members of the public were not involved in the design, conduct, reporting or dissemination plans of our research.

## Abbreviations

AI: artificial intelligence
API: application programming interface
CIViC: Clinical Interpretation of Variations in Cancer
CUP: cancer of unknown primary
DOID: disease ontology ID
HGVS: Human Genome Variation Society
HRD: homologous recombination deficiency
LLMs: large language models
mCRPC: metastatic castration-resistant prostate cancer
MMR: mismatch repair
MONDO: Monarch Disease Ontology
MSI: microsatellite instability
MTB: molecular tumour boards
NCIt: National Cancer Institute thesaurus
NER: named entity recognition
NGS: next-generation sequencing
NLP: natural language processing
NSCLC: non-small cell lung cancer
OBI: Ontology of Biomedical Investigations
OCR: optical character recognition
PC: prostate cancer
RAG: retrieval-augmented generation
RWE: real-world evidence
SNP: single-nucleotide polymorphism
TKIs: tyrosine kinase inhibitors
TMB: tumour mutational burden
UMAP: uniform manifold approximation and projection
VUS: variant of uncertain significance

