## Supplementary file for "Variantscape: Retrospective Study of Large Language Model-Based Extraction of Variant-Treatment-Cancer Associations from Published Oncology Abstracts"

### SUPPLEMENTARY MATERIAL

Marie Wosny[1,2], Andrei Stefan Blindu[3,4,5], Maximilian Boesch[6,7,8], Tobias Peres[6], Thibault Niederhauser[9], Martin Früh[6,10], Christian Rothermundt[11], Janna Hastings[3,12] *

[1] School of Medicine, University of St.Gallen (HSG), St Gallen, Switzerland

[2] Institute for Implementation Science in Health Care, University of Zurich (UZH), Zurich, Switzerland
[3] Idiap Research Institute, Switzerland

[4] Department of Electrical, Computer and Biomedical Engineering, University of Pavia, Via Ferrata, 5, 27100 Pavia, Italy

[5] enGenome s.r.l., Via Ferrata, 5, 27100 Pavia, Italy

[6] HOCH Health Ostschweiz, Kantonsspital St.Gallen, Universitäres Lehr- und Forschungsspital, Department of Medical Oncology and Hematology, St.Gallen, Switzerland

[7] HOCH Health Ostschweiz, Kantonsspital St.Gallen, Universitäres Lehr- und Forschungsspital, Lung Center, St.Gallen, Switzerland

[8] Medical Faculty, University of Basel, Basel, Switzerland

[9] Department of Quantitative Biomedicine, University of Zurich (UZH), Zurich, Switzerland

[10] Department of Oncology, Inselspital Bern, Bern, Switzerland

[11] Department of Medical Oncology and Cancer Center, Luzerner Kantonsspital (LUKS), Lucerne, Switzerland

[12] Swiss Institute of Bioinformatics (SIB), Lausanne, Switzerland

### Table of supplementary tables

Supplementary table 1: Performance evaluation of cancer type extraction. 3

Supplementary table 2: Performance evaluation of treatment extraction. 3

Supplementary table 3: Performance evaluation of large language model prompts for study design classification. 4

Supplementary table 4: Evaluation of literature-derived variant-cancer-treatment associations. 5

Supplementary table 5: Comparative overview of cancer variant resources and the Variantscape platform. 14

### Table of supplementary figures

Supplementary figure 1: Temporal analysis of biomedical publications (2014-2026) from OpenAlex: yearly, monthly, and weekly trends. 6

Supplementary figure 2: Distribution of the number of gene mentions per biomedical publication (n=886,195). 7

Supplementary figure 3: Distribution of study design types across 37,895 articles classified by DeepSeek-V4.1-Flash. 7

Supplementary figure 4: Distribution of the most frequently mentioned treatments identified in the 365,742 articles (41,27%) containing treatments of the total 886,195 publications screened. 8

Supplementary figure 5: Variant-associated extraction of oncology genes. 9

Supplementary figure 6: Progressive filtering from 886,195 articles mentioning genes to the analysis dataset of 37,895 articles (2.2%) that mention a gene, a cancer type, a treatment and a variant. 10

Supplementary figure 7: Most mentioned treatment types in the final analysis dataset of 37,895 articles. 10

Supplementary figure 8: Most mentioned cancer types in the final analysis dataset of 37,895 articles. 11

Supplementary figure 9: Heatmap illustrating co-associations of variants and cancers. 12

Supplementary figure 10: Heatmap illustrating co-associations of cancers and treatments.13

*Supplementary table 1: Performance evaluation of cancer type extraction.*

| Approach/model | Precision | Recall | Accuracy | F1 Score |
|---|---|---|---|---|
| SciSpaCy en_ner_bionlp13cg_md and en_ner_bc5cdr_md combined | 0.96 | 0.75 | 0.72 | 0.84 |
| SciSpaCy en_ner_bionlp13cg_md | 0.97 | 0.75 | 0.71 | 0.84 |
| SciSpaCy en_ner_bc5cdr_md | 0.95 | 0.39 | 0.47 | 0.55 |

*Supplementary table 2: Performance evaluation of treatment extraction.*

| Approach/model | Precision | Recall | Accuracy | F1 Score |
|---|---|---|---|---|
| Ontology-guided NER | 0.99 | 0.74 | 0.73 | 0.85 |
| SciSpaCy en_ner_bionlp13cg_md and en_ner_bc5cdr_md | 0.18 | 0.36 | 0.15 | 0.24 |
| BioBERT alvaroalon2/biobert_chemical_ner | 0.12 | 0.26 | 0.27 | 0.17 |

*Supplementary table 3: Performance evaluation of large language model prompts for study design classification.*

| Prompt number | Prompt Text | Precision | Recall | Accuracy | F1 Score |
|---|---|---|---|---|---|
| Prompt1 | INSTRUCTION: You are a helpful classifier. You are given the abstract of a scientific, biomedical publication and you have to select the correct of the possible categories. The topic of the classification is '[TOPIC]'. The allowed categories are '[CATEGORIES]'. QUESTION: The abstract to be classified is '[TEXT]'. ANSWER: The correct category for this abstract is "". | 0.93 | 0.90 | 0.90 | 0.90 |
| Prompt2 | INSTRUCTION: You are a classifier for biomedical abstracts. Classify the given abstract under one of the predefined categories. CONTEXT: The classification topic is '[TOPIC]'. Allowed categories: [CATEGORIES]. QUESTION: The abstract to be classified is '[TEXT]'. REQUIREMENTS: - Select only one category from the list. - Do not create new categories. ANSWER: The correct category for this abstract is "". | 0.94 | 0.92 | 0.92 | 0.92 |
| Prompt3 | INSTRUCTION: You are a classifier for biomedical abstracts. Classify the given abstract under one of the predefined categories. Topic: '[TOPIC]'. Allowed categories: [CATEGORIES]. Abstract: '[TEXT]'. Select only one category. Do not create new categories. ANSWER: The correct category is "". | 0.93 | 0.92 | 0.92 | 0.92 |
| Prompt4 | INSTRUCTION: You are a helpful classifier. You are given the abstract of a scientific, biomedical publication and you have to select the correct of the possible study categories. The topic of the classification is '[TOPIC]'. The allowed categories are '[CATEGORIES]'. QUESTION: The abstract to be classified is '[TEXT]'. ANSWER: The correct category for this abstract is "". | 0.95 | 0.92 | 0.92 | 0.92 |

*Supplementary table 4*: Evaluation of literature-derived variant–cancer–treatment associations.

### A: Prompt used for literature-derived association verification:

```
You check associations that were automatically mined from a biomedical publication. For each numbered candidate, decide whether the title or abstract itself REPORTS that association (a finding of the study or a statement made by the authors), not merely mentions the entities.

Title: {title}
Abstract: {abstract}

Candidates:
1. variant: {variant} | cancer: {cancer} | treatment: {treatment}
2. variant: {variant} | cancer: {cancer}
...

Relations (judge the candidate's variant as a biomarker for the treatment):
- Sensitive: patients, models or cells with the variant respond to or benefit from the treatment in this cancer
- Resistant: the variant is reported to cause or predict resistance, reduced response or shorter benefit to the treatment in this cancer, e.g. a mutation in the drug target, a variant acquired on treatment that causes resistance, or cells/patients with the variant that keep growing or progress on the treatment. Do NOT use Resistant for a variant only because patients who carry it later develop resistance through a different alteration or mechanism; for that variant use Sensitive if a response is reported, otherwise Reported
- No response: the treatment is reported as not effective for this variant in this cancer
- Diagnostic: the variant is reported as diagnostic for this cancer
- Prognostic: the variant is reported as prognostic in this cancer
- Reported: the association is reported, but none of the above applies
- Not reported: the association is not reported (only co-mentioned, mentioned in background, a different variant/cancer/treatment, or an entity that is wrong for this text)

For candidates without a treatment, judge the variant-cancer association only.
Example: in "NRAS mutations caused acquired resistance to vemurafenib in BRAF V600E melanoma", NRAS with vemurafenib is Resistant, BRAF V600E with vemurafenib is not (Sensitive or Reported).
Answer with only a JSON array, one object per candidate:
[{"id": 1, "relation": "<relation>", "quote": "<shortest verbatim excerpt from the title or abstract that supports it; empty for Not reported>"}]
```

**B. Recall of known associations** (literature evidence only; the curated CIViC tier is excluded)

| Reference set | Associations (n) | Recall, exact | Recall, exact or related | Recall, reachable | Direction agreement |
|---|---|---|---|---|---|
| CIViC, all levels | 1,426 | 0.44 | 0.54 | 0.68 | 0.87 |
| CIViC, Tier I-A assertions | 25 | 0.92 | 0.96 | 1.00 | 1.00 |
| CIViC, level A evidence | 71 | 0.76 | 0.82 | 0.91 | 1.00 |
| MOAlmanac, all levels | 384 | 0.45 | 0.61 | 0.70 | 0.92 |
| MOAlmanac, FDA-approved | 163 | 0.64 | 0.89 | 0.95 | 0.98 |

**Exact:** the variant–cancer–treatment triple was verified in at least one abstract.
**Related:** the triple was verified for a parent or subtype cancer type in OncoTree, or for a variant at the same codon.

**Reachable:** only associations whose variant and treatment occur in the literature-derived graph are counted (CIViC n = 1,134; MOAlmanac n = 336).
**Direction agreement:** whether the consensus relation (Sensitive vs Resistant, Reduced sensitivity or No response) matches the reference, counted only for found associations where both sides have a direction. A further 5.2% (CIViC) and 2.3% (MOAlmanac) of these associations were "Mixed": the abstracts disagree and no relation reaches a 60% majority.
**Literature-only:** verified associations that match no reference association.

**C. Precision of literature-only associations** (manual annotation)

| **Papers supporting the association** | **Associations (n)** | **Sampled** | **Annotator 1** | **Annotator 2** | **Both annotators** |
|---|---|---|---|---|---|
| 1 | 17,537 | 20 | 0.85 | 0.80 | 0.70 |
| 2–3 | 3,905 | 20 | 0.94 | 0.90 | 0.89 |
| ≥4 | 1,639 | 20 | 0.75 | 0.85 | 0.70 |
| Overall (weighted by stratum size) | 23,081 | 60 | 0.86 | 0.82 | 0.73 |

**Sample:** 20 drawn at random per stratum; Annotator 1 judged 18 in the 2–3 stratum.
**Both annotators:** neither annotator judged the association unsupported.

Inter-annotator agreement on whether an association was supported 81% (n = 58; Cohen's κ = 0.24, Gwet's AC1 = 0.75).

*Supplementary figure 1: Temporal analysis of biomedical publications (2014-2026) from OpenAlex: yearly, monthly, and weekly trends.*

Articles dated 1 January (11.4%) mostly have only a publication year recorded; weekly counts are shown as a 4-week rolling mean, and 2026 is partial (until 18 October).

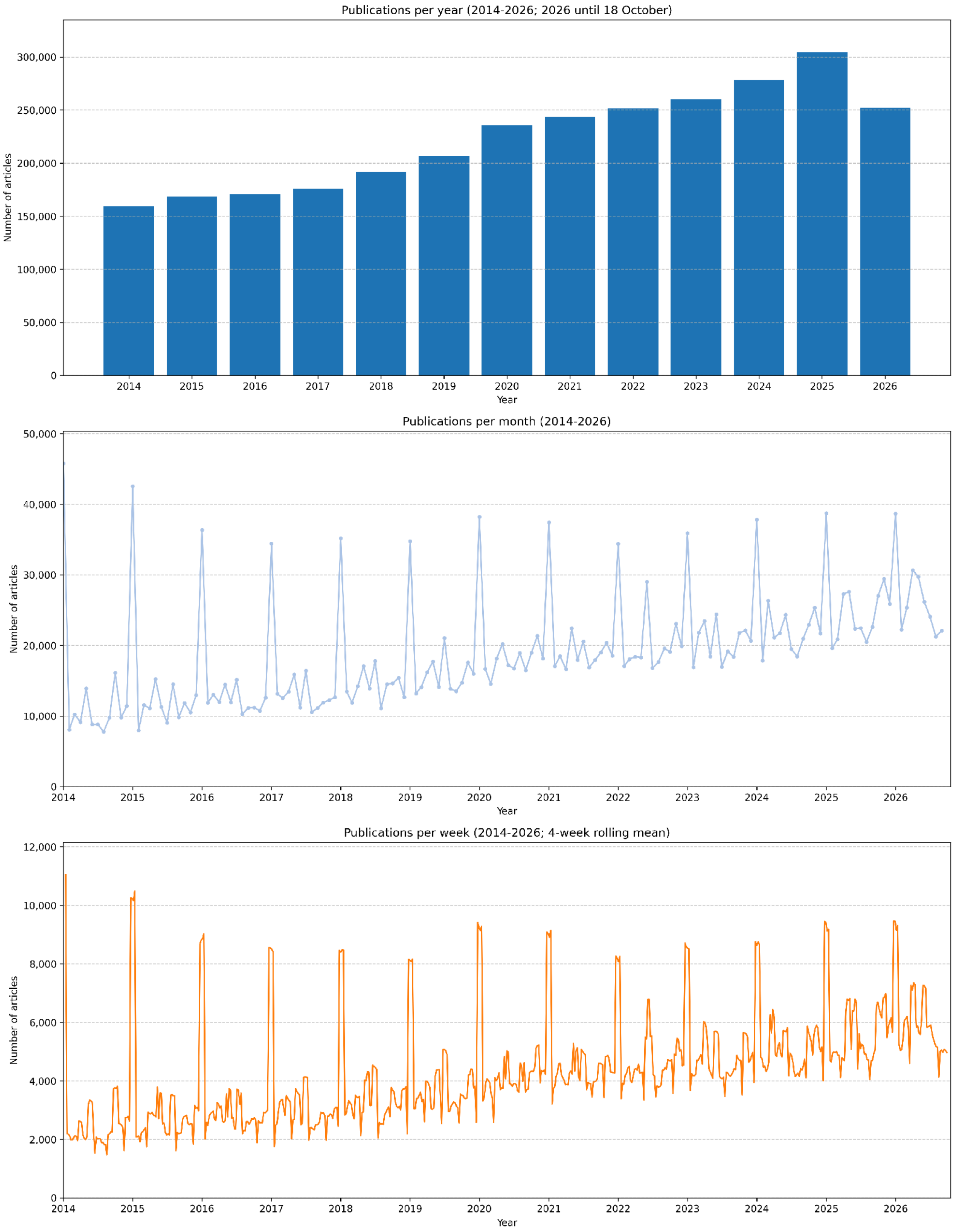

*Supplementary figure 2: Distribution of the number of gene mentions per biomedical publication (n = 886,195).*

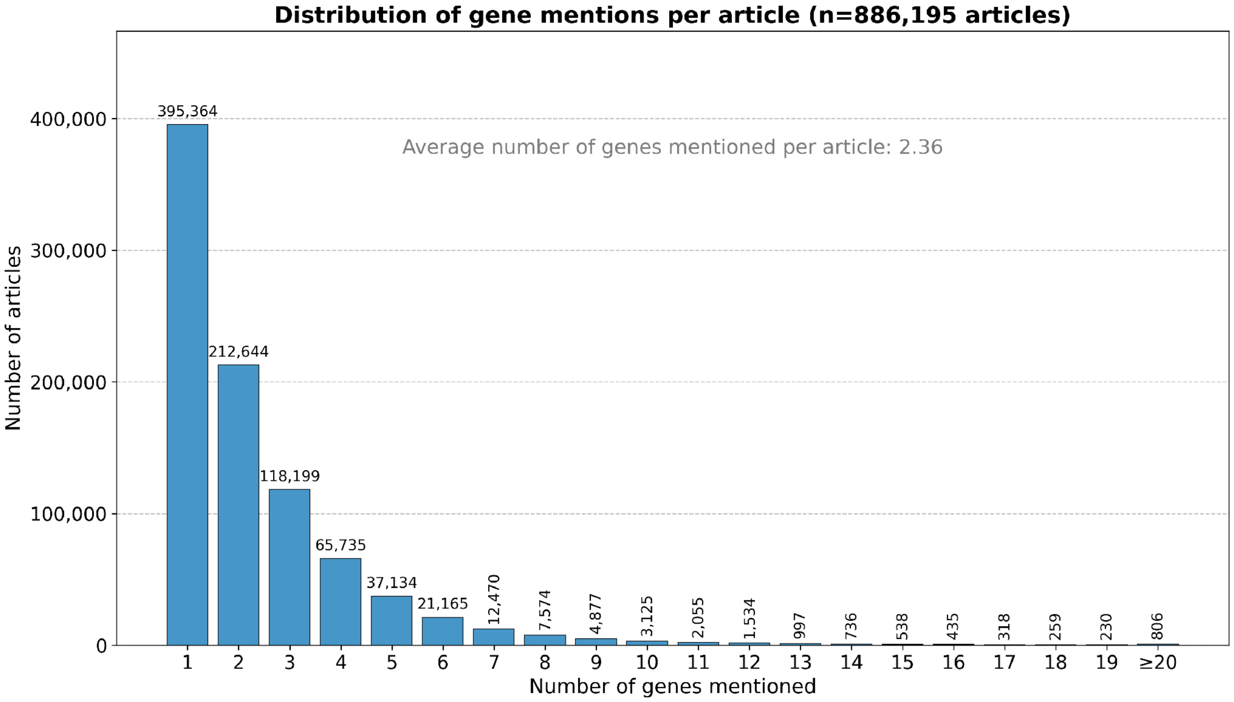


*Supplementary figure 3: Distribution of study design types across 37,895 articles classified by DeepSeek-V4.1-Flash*

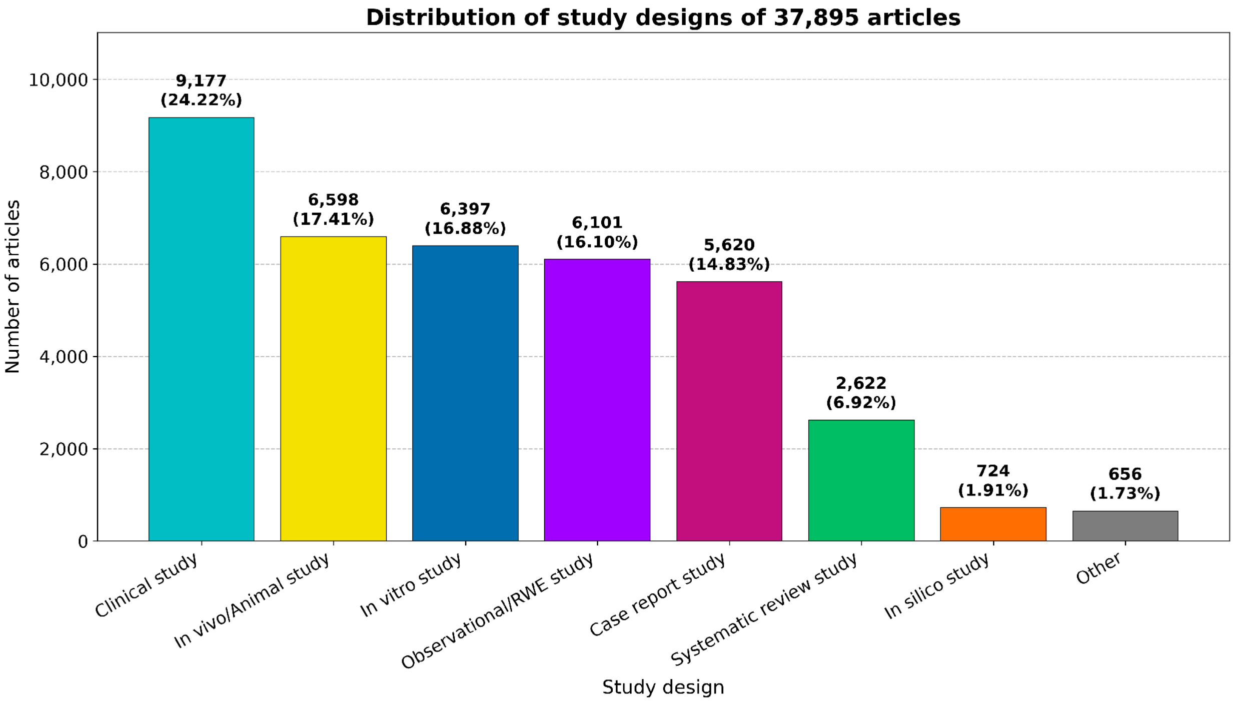

*Supplementary figure 4: Distribution of the most frequently mentioned treatments identified in the 365,742 articles (41.27%) containing treatments, of the total 886,195 publications screened.*

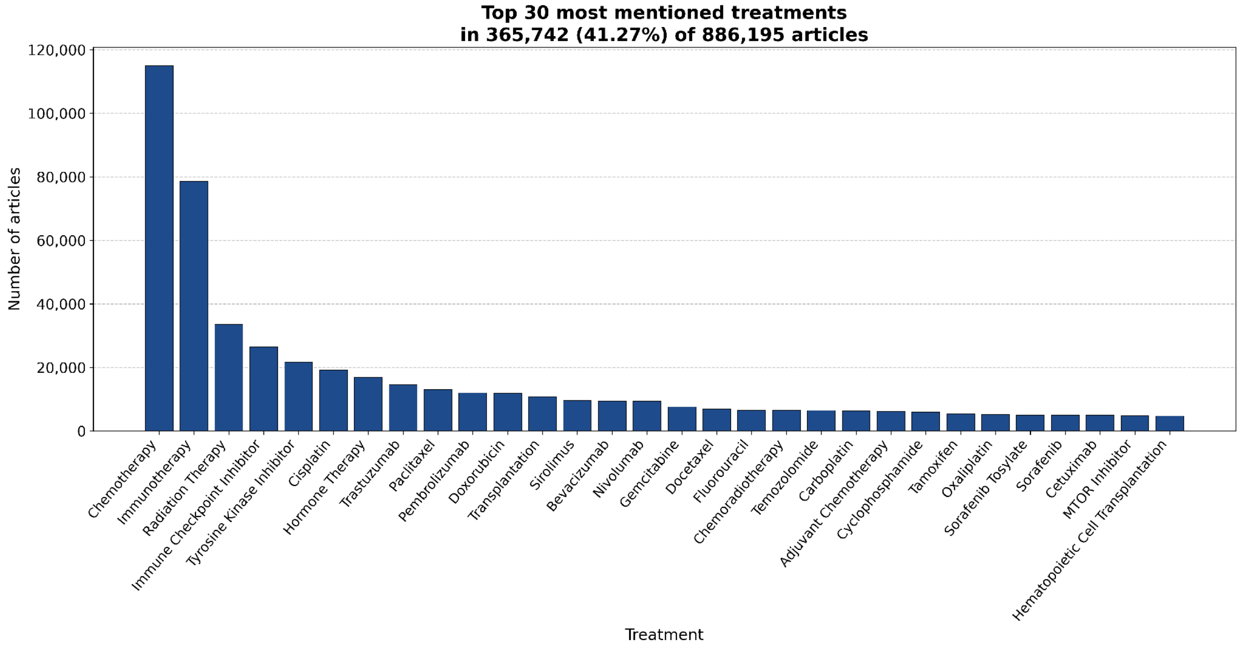


*Supplementary figure 5: Variant-associated extraction of oncology genes.*

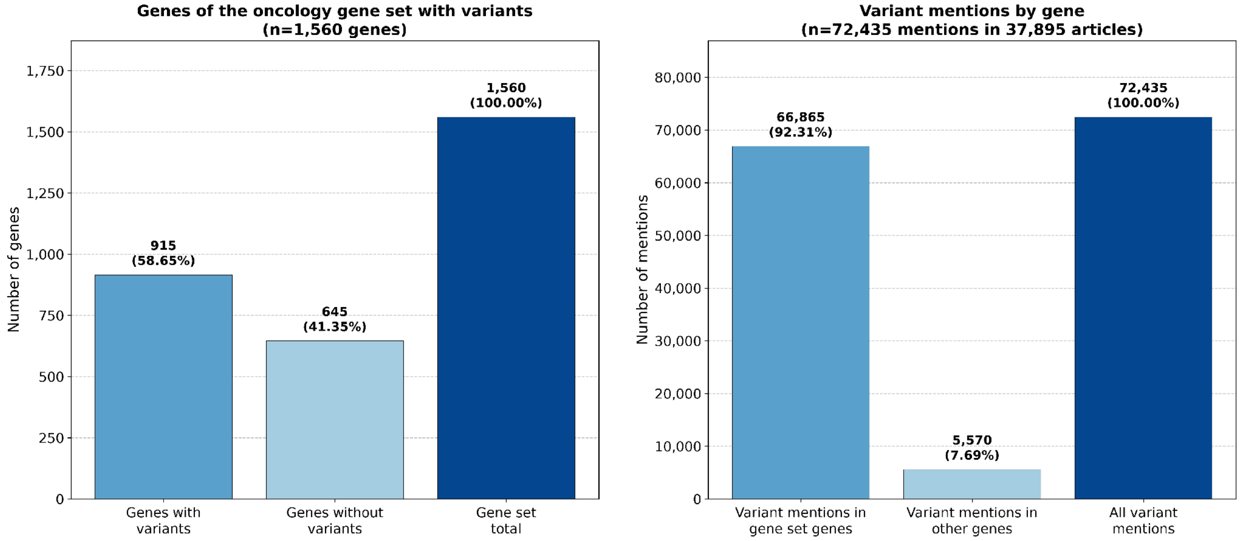

*Supplementary figure 6: Progressive filtering from 886,195 articles mentioning genes of the oncology gene set to the analysis dataset of 37,895 articles (2.2% of the original dataset) that mention a gene, a cancer type, a treatment and a variant.*

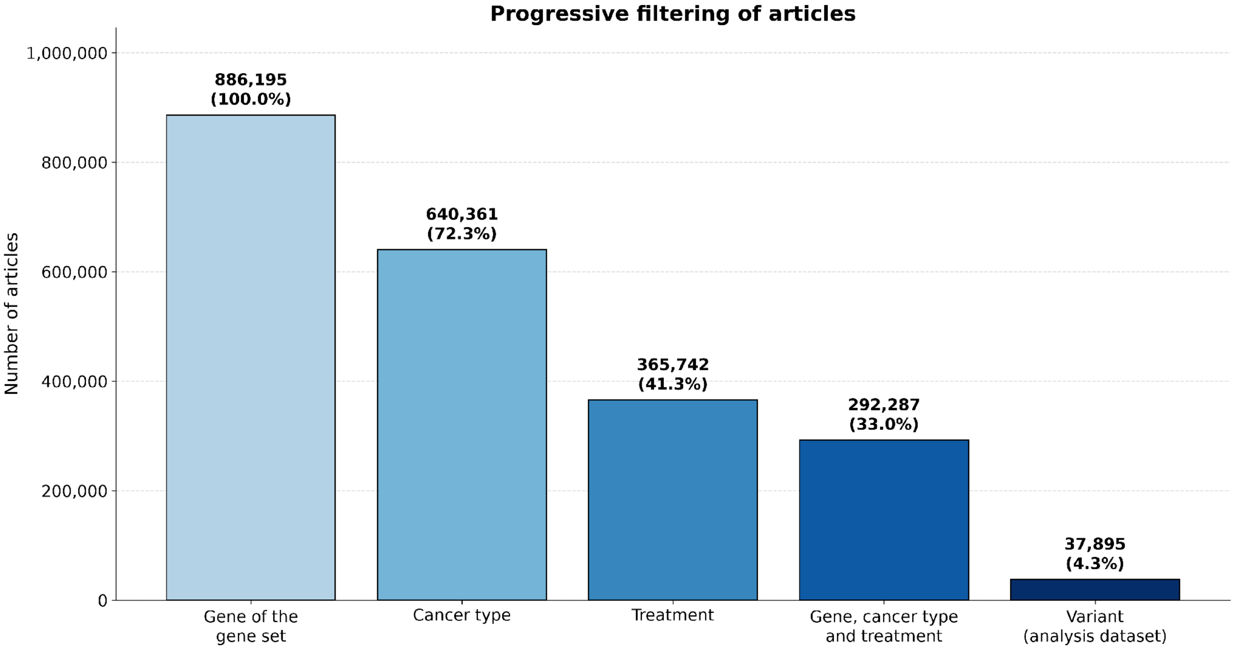


*Supplementary figure 7: Most mentioned treatment types in the final analysis dataset of 37,895 articles.*

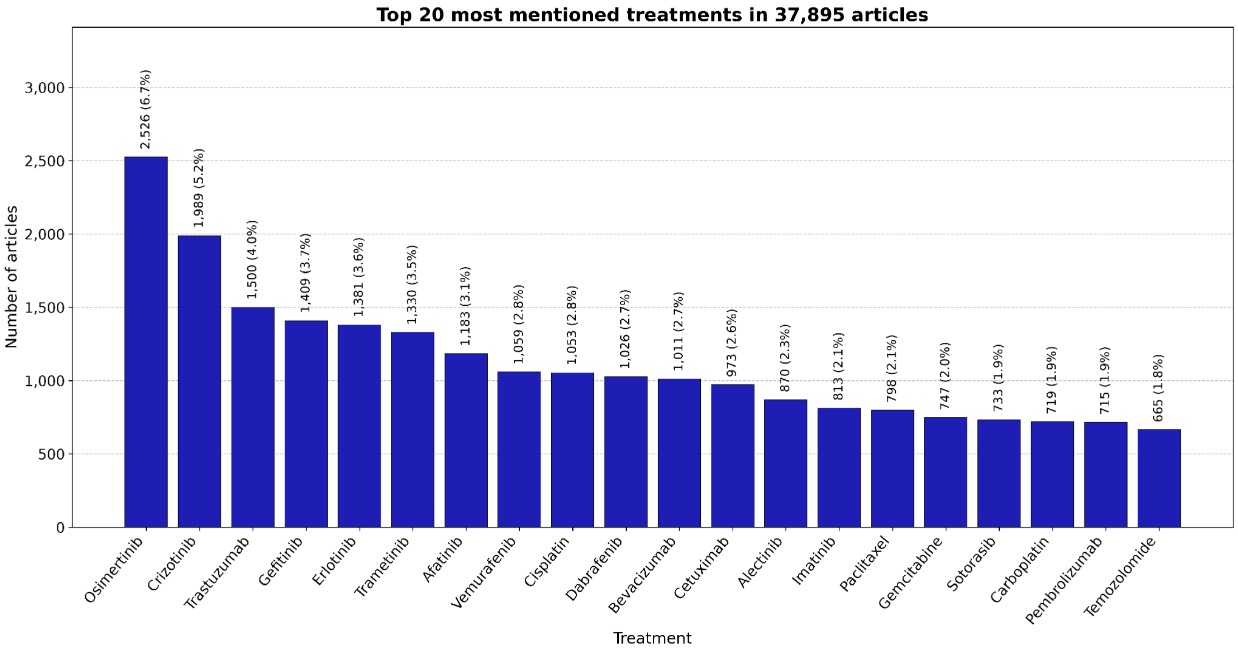

*Supplementary figure 8: Most mentioned cancer types in the final analysis dataset of 37,895 articles.*

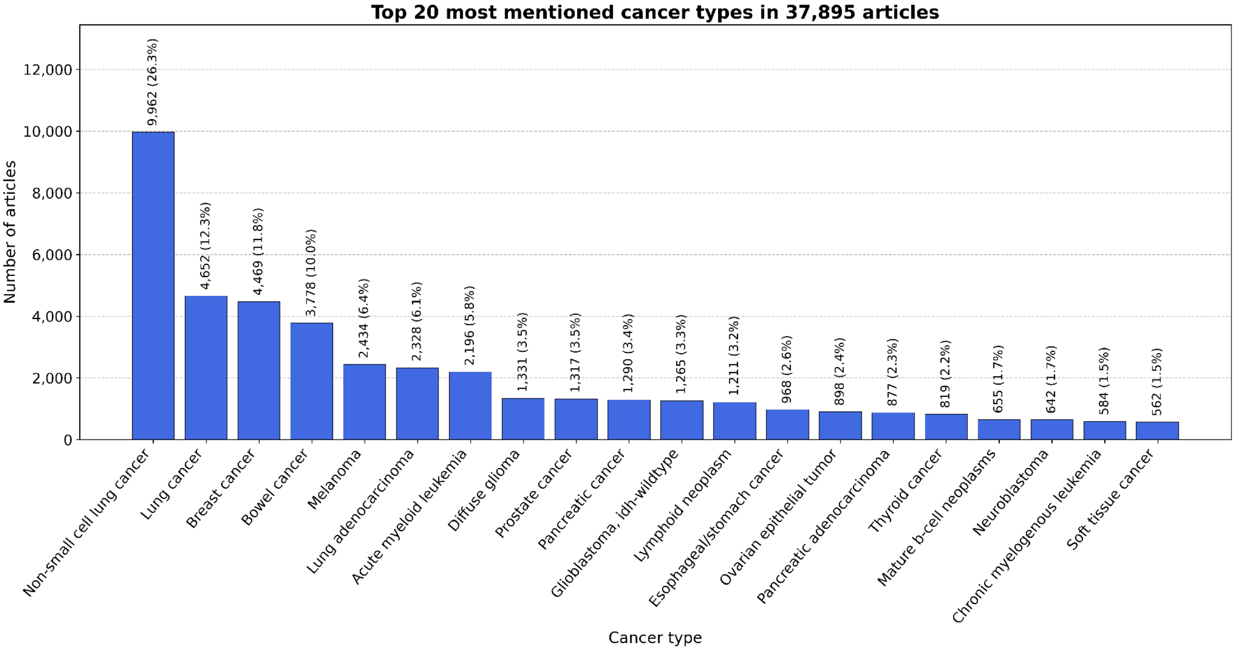

***Supplementary figure 9:*** *Heatmap of verified cancer–variant associations: share of the articles of each cancer type (40 most frequent; number of articles in brackets) in which the association with each variant (40 most frequent) was verified.*

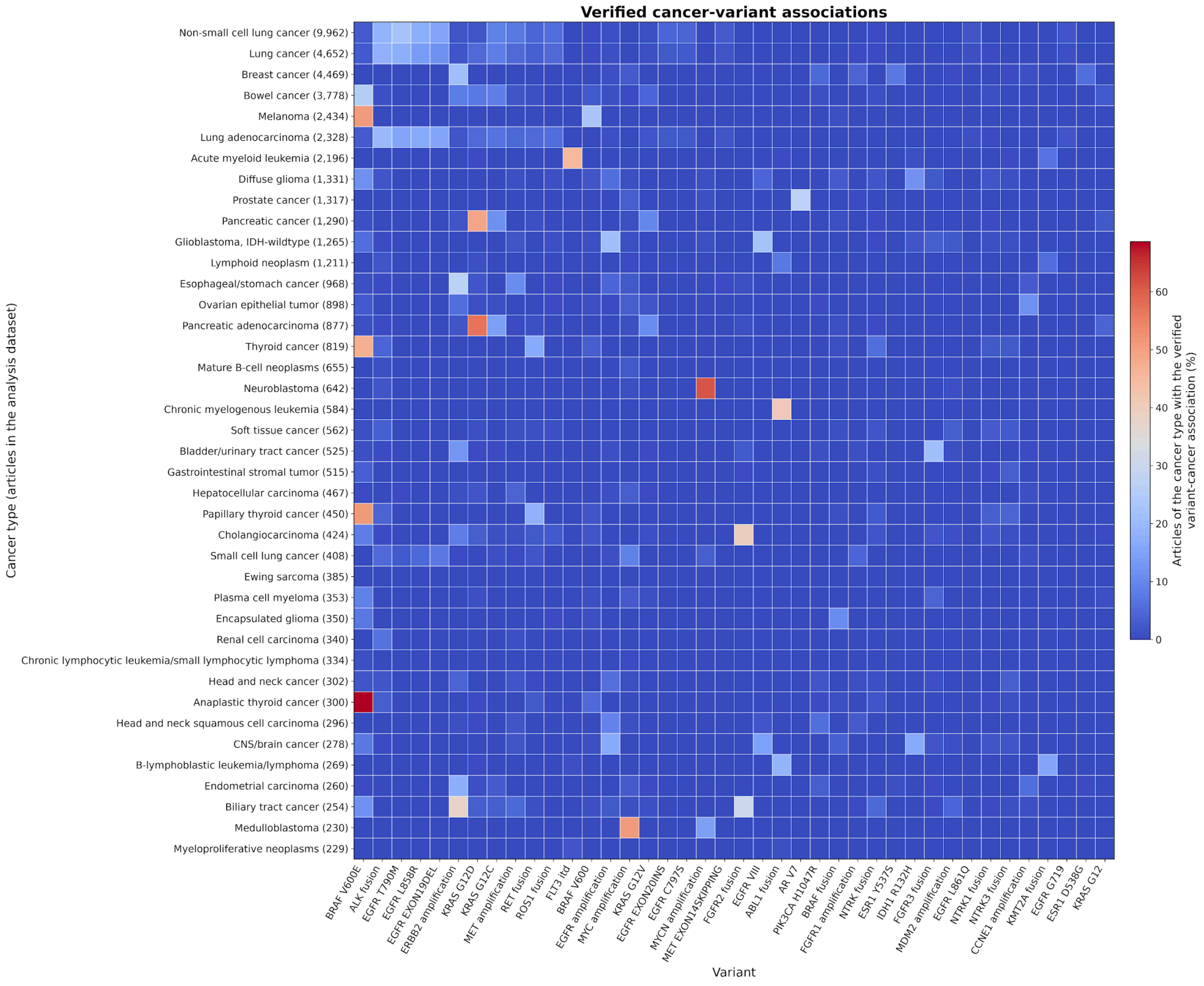

***Supplementary figure 10:*** *Heatmap illustrating verified co-associations of cancers and treatments: share of the articles of each cancer type (40 most frequent; number of articles in brackets) in which the association with each treatment (40 verified in most articles; general treatment classes excluded) was verified.*

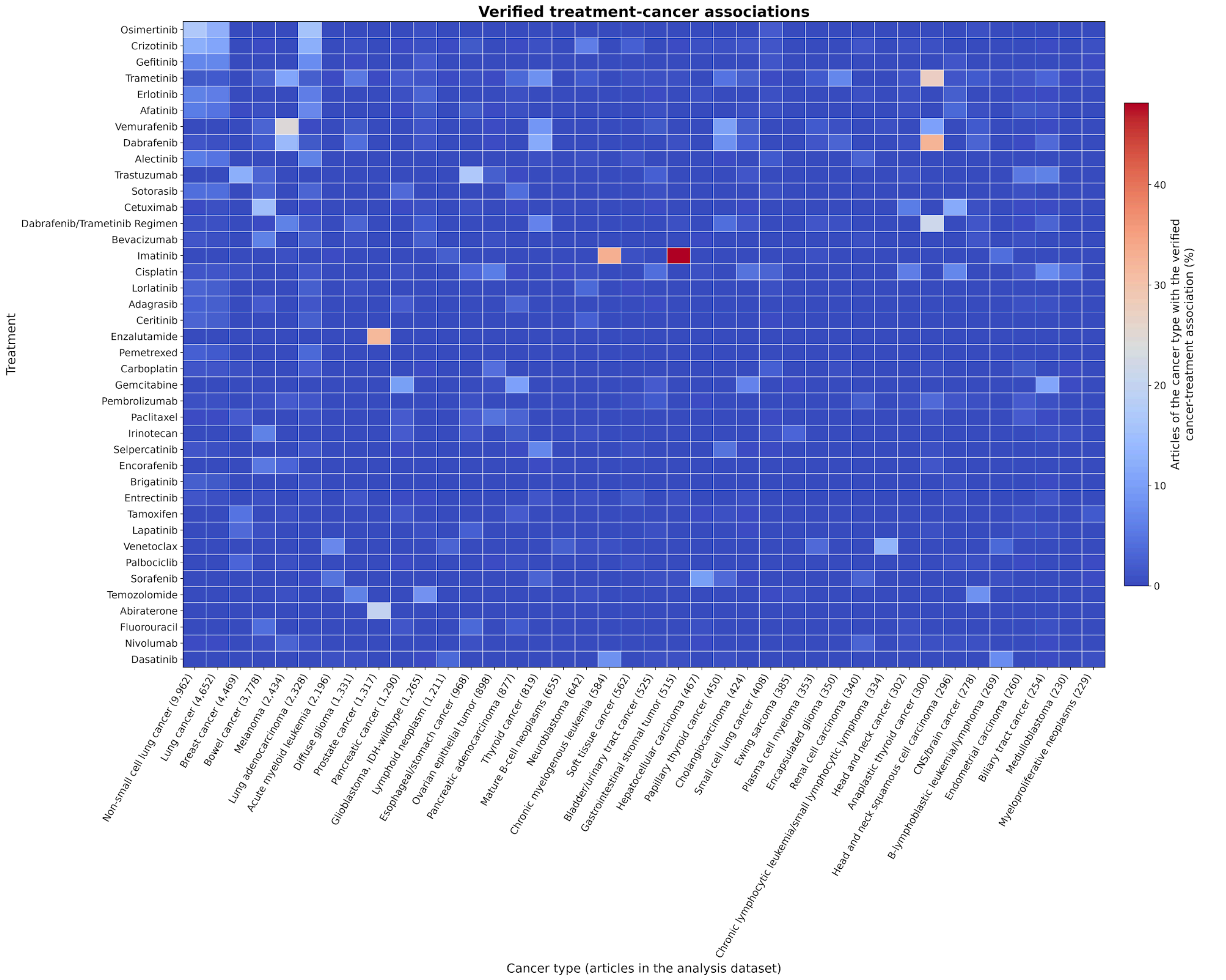

***Supplementary table 5:*** *Comparative overview of cancer variant resources and the Variantscape platform.*

| Feature | OncoKB | CIViC | ClinVar | COSMIC | VarSome | tmVar | PubTator | GenomeNexus | PeCanPIE | Variantscape |
|---|---|---|---|---|---|---|---|---|---|---|
| **Database characteristics** | | | | | | | | | | |
| **Abbreviation / meaning** | Oncology knowledge base | Clinical Interpretation of Variants in Cancer | Clinical Variant database | Catalogue Of Somatic Mutations In Cancer | NA | Text Mining for Variants | NA | NA | Pediatric Cancer variant Pathogenicity Information Exchange | Variant landscape explorer |
| **Goal / intention** | Linking variants to therapies | Interpretation of cancer-related variants with evidence levels | Aggregate pathogenicity assertions from clinical labs | Catalog somatic mutations in cancer from sequencing data | Aggregate and interpret genetic variants from multiple sources | Extract variants from text using NLP techniques | Tag biomedical concepts (genes, variants, etc.) in publications | Aggregate annotations and variant information from multiple tools for consistent output | Annotates and ranks variants by putative pathogenicity, display in an interactive web interface | Build networks of gene, variant, disease, and treatment relationships from biomedical literature |
| **URL** | https://www.oncokb.org | https://civicdb.org | https://www.ncbi.nlm.nih.gov/clinvar | https://cancer.sanger.ac.uk/cosmic | https://varsome.com | https://www.ncbi.nlm.nih.gov/research/bionlp/Tools/tmvar/ | https://www.ncbi.nlm.nih.gov/research/pubtator/ | https://www.genomenexus.org | https://pecan.stjude.cloud | https://evidencedb.hastingslab.org/variantscape |
| **Sources and access** | | | | | | | | | | |
| **Source / curation** | Structured (expert curation from clinical guidelines/trials) | Structured (crowdsourced evidence + expert-reviewed) | Structured (aggregated clinical submissions with expert/community assertions) | Structured (manual literature + project-based somatic curation by experts) | Aggregated (databases + computational predictions; automated NLP) | Unstructured (text-mined variant mentions; automated NLP) | Unstructured (NER-tagged biomedical concepts from abstracts; automated NLP) | Aggregated (harmonized structured data; automated tool aggregation) | Mixed (structured databases, literature, rule-based scoring; automated pipeline) | Unstructured (title + abstracts with graph structure using NLP + LLMs) |
| **Cost** | Free for academics | Free | Free | Free for academic, non-commercial use (paid for bulk) | Partially free (paid for bulk) | Free | Free | Free | Free | Free |
| **Methods and clinical relevance** | | | | | | | | | | |
| **Literature mining** | No | Manual curation | No | No | Yes (variant mentions) | Yes (extracted variant mentions) | Yes (NER + relation extraction) | No | Yes (mines PubMed, etc.) | Yes, with RAG-based literature search for top 5 associated papers |
| **AI / LLM integration** | No | No | No | No | No | No | No | No | Partial (heuristics) | Yes (LLMs + NLP), LLM-based semantic interpretation |
| **Clinical associations** | Strong | Strong | Limited | Limited | No | No | Limited (indirect) | Strong | Strong | Strong (treatment nodes) |
| **Clinical relevance scoring** | Yes (manual) | Yes (manual) | Yes (pathogenicity) | No | No | No | No | Limited | Yes | Yes (automated) |
